# Individual-First Brain State Modeling Reconciles Personalized Prediction with Generalizable Mechanistic Interpretation in Precision Psychiatry

**DOI:** 10.64898/2026.04.28.26351834

**Authors:** Chen Ran, Xinyun Li, Chenfei Ye, Ting Ma

## Abstract

Neuropsychiatric heterogeneity creates a fundamental trade-off between personalized prediction and generalizable mechanistic interpretation. Conventional brain metastate models enforce shared state repertoires, inherently suppressing clinically meaningful individual variation, whereas individualized brain states lack direct correspondence across participants, limiting cross-subject comparison. We present Individual-specific Brain State Space (IBSS), an interpretable individual-first framework that estimates brain states separately for each participant and models their spatiotemporal organization for individualized prediction. We then cluster similar individualized states across participants into shared metastates, thereby reconstructing a unified metastate space for generalizable neurodynamic interpretation. Across four large-scale fMRI cohorts (HCP-task, ABIDE-I/II, and DIRECT-II), the IBSS-based graph attention network achieved state-of-the-art performance in cognitive decoding and ASD/MDD identification, yielding accuracy gains of 16.66–36.69% over conventional group-first metastate approaches. These gains were primarily attributable to individualized state configurations rather than model complexity. Metastate reconstruction further revealed reproducible, symptom-linked neurodynamic patterns, including frontoparietal control network hyper-engagement in ASD and distinct default mode–limbic dynamics related to depressive and anxiety symptoms in MDD. This paradigm bridges personalized diagnostic prediction and shared neurodynamic interpretation, enabling robust heterogeneity characterization for precision psychiatry.

## Introduction

Neuropsychiatric disorders are defined by profound biological and clinical heterogeneity, creating a persistent tension between personalized predictive modeling and generalizable mechanistic insight (1). Accurate individual-level assessment requires neural representations that preserve each person’s unique functional architecture, whereas mechanistic inference requires a shared reference framework to compare neurobiological alterations across individuals (2). This trade-off is particularly challenging in autism spectrum disorder (ASD) and major depressive disorder (MDD), where patients with the same diagnosis may differ markedly in neural organization, clinical manifestations, developmental trajectories, and treatment responses (3–9). Group-averaged analyses risk erasing clinically meaningful heterogeneity, whereas purely individualized representations hinder the identification of generalizable neurobiological patterns shared across patients. A central unresolved challenge is thus to retain individual-specific neural information without sacrificing cross-subject interpretability.

Brain state analysis provides a natural framework in which individualized neural dynamics and shared functional organization can be jointly investigated. By framing large-scale neural activity as transitions among a shared repertoire of recurring brain states, these methods establish a common functional reference across individuals while enabling the characterization of subject-specific temporal trajectories (10–13). Dynamic properties derived from these trajectories, such as state fractional occupancy, dwell time, and transition probability, have provided valuable insights into the neural mechanisms underlying cognition and neuropsychiatric disorders (14–19). In particular, altered brain state dynamics have been increasingly implicated in ASD (20, 21) and MDD (22–24), suggesting that dynamic neural organization may offer clinically relevant information beyond conventional static functional connectivity. Despite this progress, translating these brain-state signatures into reliable individualized predictions remains a fundamental bottleneck.

A core limitation lies in the group-first paradigm underlying most conventional brain state identification methods. These approaches first impose a single, shared state repertoire across individuals, prioritizing group-level commonality over individual specificity and restricting inter-individual variation primarily to state occurrence and transition dynamics (10, 12–25). While this design simplifies group comparisons, it inherently suppresses individualized variation in the functional organization of brain states. Under this formulation, spatial state configurations are fixed at the group level, while individualized temporal dynamics are typically reduced into a limited set of handcrafted metrics. This artificially separates the two intrinsically coupled dimensions, overlooking the spatiotemporal structures of brain dynamics that encode more phenotype-linked neural features (26–29). Therefore, a framework that preserves individualized brain state organization while jointly modeling its spatial and temporal structure is needed to fully capture clinically relevant heterogeneity for individual-level psychiatric prediction.

Identifying brain states independently for each participant offers a conceptually straightforward means of preserving such heterogeneity (30). Individualized states can capture participant-specific activation patterns and transition dynamics that are obscured by group-level modeling, thereby providing richer neural representations for personalized prediction. Nevertheless, independently estimated brain states lack explicit correspondence across individuals: a state identified in one participant does not necessarily have a direct counterpart in another (31). Individualization therefore leaves the other side of the trade-off unresolved, because individualized states cannot be directly compared or readily summarized across participants. Existing approaches thus face a fundamental tension between preserving inter-individual neurobiological heterogeneity and establishing a shared reference for generalizable mechanistic interpretation (32). Resolving this tension requires a framework that does not impose a common state repertoire before individual variation is modeled, but instead reconstructs shared dynamic organization from individualized brain states (33).

To address this gap, we propose Individual-specific Brain State Space (IBSS), an individual-first brain-state framework that decouples individual state estimation from cross-subject alignment. Specifically, we first construct a spatiotemporal representation of individualized brain states for each participant and subsequently recover the unified metastate space underlying these individualized states (Figure 1). For each participant, brain states are independently inferred using a hidden Markov model and represented as a directed graph, in which nodes encode individualized whole-brain activation patterns and edges capture transitions between states. We then introduce an IBSS-based graph attention network (IBSS-GAT) to learn participant-level representations from the spatiotemporal organization of each IBSS for cognitive decoding and disease identification. Rather than treating these individualized representations as isolated predictive features, we organize similar states across participants into shared metastates, enabling their mechanistic interpretation through cross-subject metastate dynamics.

**Figure 1.**
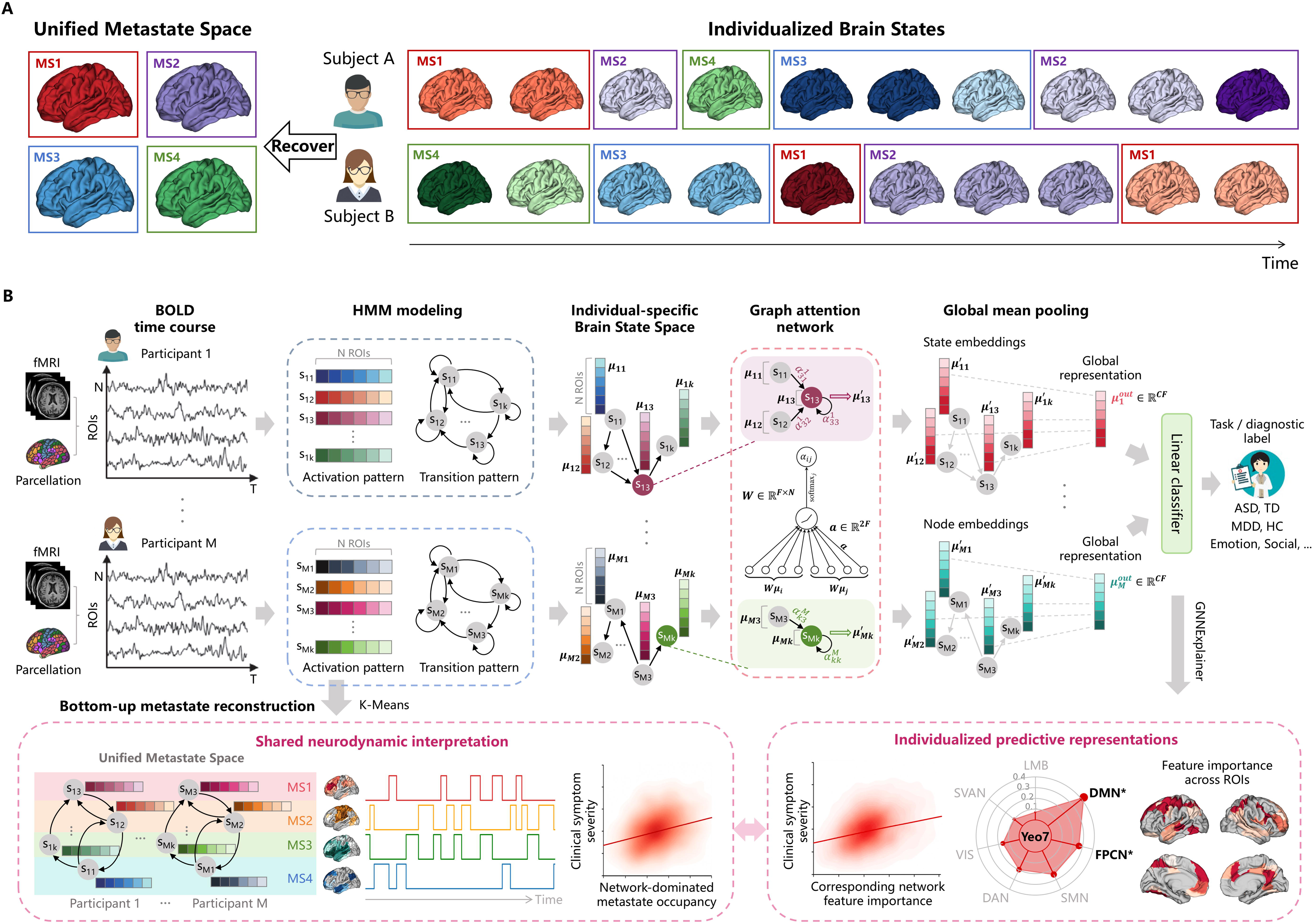
Conceptual overview and analytical framework of IBSS. **(A)** Individual-first, group-reconstruction strategy. Brain states are independently estimated for each participant, allowing their number, activation patterns, and temporal organization to vary across individuals. States with similar activation patterns are subsequently grouped to recover a unified metastate space, establishing cross-subject correspondence without imposing shared states a priori. Colors indicate metastate assignments. **(B)** Analytical workflow. Regional BOLD time courses were extracted from parcellated fMRI data and modeled using individualized hidden Markov models. Each participant’s individual-specific brain state space was represented as a directed graph, with states as nodes, activation patterns as node features, and transitions as edges. A graph attention network generated participant-level representations for cognitive decoding or psychiatric-disorder identification, while GNNExplainer quantified predictive feature importance. In parallel, individualized states were clustered to reconstruct a unified metastate space, enabling analyses of metastate dynamics and their clinical associations. Together, these two branches link individualized predictive representations to shared neurodynamic interpretation. BOLD, blood-oxygen-level-dependent; HMM, hidden Markov model; ROI, region of interest; ASD, autism spectrum disorder; TD, typically developing; MDD, major depressive disorder; HC, healthy control.

We evaluated the proposed framework across four large-scale neuroimaging cohorts spanning task-evoked cognition (HCP-task), ASD (ABIDE-I), and MDD (DIRECT-II), with the complete analytical pipeline independently replicated in ABIDE-II. IBSS-GAT achieved state-of-the-art performance across all tasks, with substantial accuracy gains over conventional group-first metastate approaches. Reconstruction of the unified metastate space further linked IBSS-derived neural representations to clinically relevant dynamic alterations. In ASD, convergent analyses revealed a reproducible frontoparietal control network (FPCN)–dominated metastate whose preferential engagement was associated with symptom severity across independent cohorts. In MDD, individualized neural representations were dynamically expressed through metastate involving the default mode network (DMN) and limbic system (LMB). Together, these findings establish an individual-first, group-reconstruction paradigm that reconciles personalized prediction with shared neurodynamic interpretation, providing a generalizable framework for studying heterogeneity in precision psychiatry.

## RESULTS

### IBSS-GAT Achieves State-of-the-Art Performance in Cognitive Decoding and Disease Identification

To evaluate whether the individual-specific brain state space (IBSS) supports individualized prediction, we first benchmarked IBSS-GAT on the well-characterized HCP-task cohort, which provides gold-standard task-evoked neural signatures with established cognitive correlates. We then evaluated its performance on two heterogeneous psychiatric cohorts, ABIDE-I (ASD) and DIRECT-II (MDD), to assess its generalizability for disease identification.

IBSS-GAT achieved state-of-the-art performance across all evaluated prediction tasks. On the HCP-task cohort, the framework decoded eight brain states (emotion, gambling, language, motor, relational, social, working memory, and resting state) with a mean accuracy (ACC) of 99.30 ± 0.42% and an area under the curve (AUC) of 99.96 ± 0.05% across 10-fold cross-validation (Table 1 and Table S4). Robust classification performance was further observed for ASD versus TD in ABIDE-I (ACC = 77.26 ± 3.70%, AUC = 78.59 ± 4.60%) and MDD versus HC in DIRECT-II (ACC = 70.68 ± 1.64%, AUC = 73.25 ± 2.50%), with consistent performance across alternative brain parcellation atlases (Table S5).

**Table 1.** Classification performance of IBSS-GAT in cognitive decoding and disease identification.

| Method |  | HCP-task<br>(8-task Classification) |  | ABIDE-I (ASD vs. TD) |  |  |  | ABIDE-II (ASD vs. TD) |  |  |  | DIRECT-II (MDD vs. HC) |  |  |  |
| --- | --- | --- | --- | --- | --- | --- | --- | --- | --- | --- | --- | --- | --- | --- | --- |
|  |  | ACC (%) | AUC (%) | ACC (%) | SEN (%) | SPE (%) | AUC (%) | ACC (%) | SEN (%) | SPE (%) | AUC (%) | ACC (%) | SEN (%) | SPE (%) | AUC (%) |
| Conventional group-first metastate approaches | FO-SVM | 61.42±1.23 | 91.65±0.59 | 51.33±2.43 | 45.37±8.04 | 57.92±10.56 | 49.52±5.87 | 51.53±4.59 | 39.55±5.22 | 62.08±7.26 | 45.91±5.20 | 55.69±3.60 | 67.43±3.87 | 42.21±4.75 | 42.70±4.93 |
|  | DT-SVM | 55.95±1.36 | 89.62±0.51 | 51.00±3.40 | 34.45±9.13 | 67.42±8.21 | 52.86±4.14 | 52.38±3.85 | 37.64±5.38 | 65.33±7.26 | 48.69±5.50 | 53.94±1.57 | 64.70±3.21 | 41.59±4.49 | 45.04±3.43 |
|  | TP- SVM | 60.66±1.85 | 91.36±0.65 | 55.41±5.09 | 47.11±6.40 | 63.62±9.49 | 41.99±4.33 | 52.81±3.50 | 41.69±9.73 | 62.62±10.74 | 47.52±5.86 | 55.94±3.86 | 70.23±3.77 | 39.55±4.98 | 43.50±5.32 |
|  | MS-GAT | 62.61±1.57 | 90.46±0.71 | 52.99±3.13 | 26.08±21.88 | 77.87±21.83 | 52.47±5.66 | 51.67±5.94 | 58.56±41.72 | 45.57±43.41 | 53.58±7.68 | 54.02±1.28 | **98.29±3.57** | 3.21±6.43 | 61.01±3.86 |
| Baseline deep learning models | BrainGNN | 97.33±0.79 | 99.82±0.14 | 70.98±4.59 | 61.89±7.49 | 79.36±9.13 | 71.83±5.77 | 71.14±3.72 | 73.08±9.20 | 69.38±9.13 | 71.57±4.37 | 65.26±2.37 | 72.33±4.28 | 57.14±7.35 | 66.94±3.33 |
|  | ST-GCN | 99.26±0.39 | 99.94±0.04 | 69.76±2.44 | 66.99±3.25 | 72.34±4.66 | 71.37±2.89 | 68.10±3.63 | 61.70±6.01 | 73.72±5.88 | 69.75±4.85 | 64.51±2.66 | 70.78±4.10 | 57.33±5.06 | 67.15±4.18 |
|  | dFCExpert | 97.61±0.67 | 99.91±0.09 | 69.09±3.22 | 60.55±14.08 | 77.02±9.41 | 72.81±4.43 | 71.43±3.98 | 71.25±7.74 | 71.58±8.42 | 71.63±5.31 | 68.64±2.77 | 72.15±9.19 | 64.85±11.07 | 70.24±2.22 |
|  | BNT | 98.91±0.41 | 99.89±0.15 | 74.06±3.24 | **74.32±6.70** | 73.83±7.62 | 72.79±3.58 | 74.60±4.85 | 70.05±10.04 | 78.60±6.65 | 73.84±6.32 | 69.22±3.01 | 75.06±4.70 | 62.52±6.60 | 71.97±3.39 |
| **Ours** | **IBSS-GAT** | **99.30±0.42** | **99.96±0.05** | **77.26±3.70** | 71.99±8.95 | **82.13±4.68** | **78.59±4.60** | **77.49±2.44** | **74.41±6.54** | **80.25±6.91** | **79.17±4.02** | **70.68±1.64** | 73.27±4.69 | **67.71±5.50** | **73.25±2.50** |

Note: All data are presented in mean ± standard deviation mode, and best performances are bolded. FO, fractional occupancy; DT, dwell time; TP, transition probability; SVM, support vector machine; BrainGNN, brain graph neural network; ST-GCN, spatio-temporal graph convolution network; dFCExpert, dynamic functional connectivity expert; BNT, brain network transformer; MS-GAT, metastate space-graph attention network; IBSS-GAT (our method), individual-specific brain state space-graph attention network; ASD, autism spectrum disorder; TD, typically developing; MDD, major depressive disorder; HC, healthy control; ACC, accuracy; SEN, sensitivity; SPE, specificity; AUC, area under curve.

To determine whether these performance gains arose from individualized brain state representations, we compared IBSS-GAT with its group-level counterpart (MS-GAT; see Methods for details). Replacing the conventional group-first metastate space with the individual-specific brain state space improved cognitive decoding accuracy by 36.69% on the HCP-task dataset, while increasing disease classification accuracy by 24.27% for ASD and 16.66% for MDD (Table 1). Moreover, compared with conventional metastate-derived dynamic features, including fractional occupancy (FO), dwell time (DT), and transition probability (TP), IBSS-GAT achieved ACC improvements of 37.88–43.35% on the HCP-task dataset, 21.85–26.26% on the ABIDE-I cohort, and 14.74–16.74% on the DIRECT-II cohort. These findings highlight that preserving individualized brain state representations, rather than enforcing a shared metastate repertoire, substantially enhances the predictive utility of brain dynamics at the individual level.

IBSS-GAT also consistently outperformed other benchmark deep learning methods, including BrainGNN, ST-GCN, dFCExpert, and Brain Network Transformer. These competing methods achieved average ACCs ranging from 97.33% to 99.26% on the HCP task cohort, 69.09% to 74.06% on the ABIDE-I cohort, and 64.51% to 69.22% on the DIRECT-II cohort (Table 1).

### Individualized Brain State Representations Underlie the Superior Performance of IBSS-GAT

To quantify the contribution of each core design element and validate the biological rationale underlying IBSS-GAT, we performed single-variable ablation analyses (Table 2). Among all ablated components, individualized state activation patterns (AP) contributed most substantially to model performance. Replacing participant-specific activation patterns derived from individual-level HMM inference with an equal number of randomly generated activation patterns reduced accuracy by 86.52% on the HCP cognitive decoding task, 25.38% on the ABIDE-I ASD classification task, and 17.24% on the DIRECT-II MDD classification task. Together with the comparison against the group-first metastate approaches (Table 1), these findings demonstrate that the superior predictive performance of IBSS-GAT is primarily driven by biologically meaningful individualized activation patterns that preserve inter-individual spatial heterogeneity in brain-state organization.

**Table 2.** Ablation study of our IBSS-GAT model.

| Model | HCP-task<br>(8-task Classification) |  | ABIDE-I (ASD vs. TD) |  |  |  | ABIDE-II (ASD vs. TD) |  |  |  | DIRECT-II (MDD vs. HC) |  |  |  |
| --- | --- | --- | --- | --- | --- | --- | --- | --- | --- | --- | --- | --- | --- | --- |
|  | ACC (%) | AUC (%) | ACC (%) | SEN (%) | SPE (%) | AUC (%) | ACC (%) | SEN (%) | SPE (%) | AUC (%) | ACC (%) | SEN (%) | SPE (%) | AUC (%) |
| w/o AP | 12.78±0.43 | 50.28±1.84 | 51.88±0.28 | 0.00±0.00 | **100.00±0.00** | 58.48±2.69 | 48.64±2.99 | 70.00±45.83 | 30.00±45.83 | 54.81±4.73 | 53.44±0.17 | **100.00±0.00** | 0.00±0.00 | 56.25±2.89 |
| w/o TP | 96.19±0.98 | 99.71±0.18 | 69.09±2.15 | 58.73±9.44 | 78.72±8.67 | 70.20±4.01 | 68.97±4.82 | 68.51±7.21 | 69.37±10.56 | 69.45±5.98 | 65.18±2.02 | 76.15±6.96 | 52.59±9.22 | 67.24±2.86 |
| w/o GAT | 98.75±0.45 | 99.88±0.08 | 73.40±3.35 | 67.17±6.04 | 79.15±8.23 | 75.01±4.00 | 73.01±3.83 | 73.72±8.02 | 72.37±6.55 | 71.68±6.54 | 68.81±2.44 | 79.11±5.23 | 56.97±9.20 | 69.75±3.65 |
| **IBSS-GAT** | **99.30±0.42** | **99.96±0.05** | **77.26±3.70** | **71.99±8.95** | 82.13±4.68 | **78.59±4.60** | **77.49±2.44** | **74.41±6.54** | **80.25±6.91** | **79.17±4.02** | **70.68±1.64** | 73.27±4.69 | **67.71±5.50** | **73.25±2.50** |

Note: All data are presented in mean ± standard deviation mode, and the best performances are highlighted in bolded. AP, activation pattern; TP, transition pattern; GAT, graph attention network; IBSS-GAT (our method), individual-specific brain state space-graph attention network.

The remaining architectural components also contributed to model performance, although to a lesser extent. Replacing the empirically derived state transition matrix with a sparsity-matched random binary matrix reduced accuracy by 3.11%, 8.17%, and 5.50% on the HCP, ABIDE-I, and DIRECT-II datasets, respectively, indicating that biologically meaningful state transition patterns (TP) provide complementary temporal information that further improves predictive performance. Similarly, replacing the GAT module with an otherwise identical GCN reduced accuracy by 0.55%, 3.86%, and 1.87%, respectively, underscoring the importance of adaptive attention mechanisms for integrating complementary spatial (AP) and temporal (TP) information encoded in the IBSS graph.

### IBSS-GAT Learns Biologically Meaningful Neural Representations

To determine whether the representations learned by IBSS-GAT capture biologically meaningful neural information, we applied GNNExplainer (34) to quantify regional feature importance underlying model predictions across the HCP-task, ABIDE-I, and DIRECT-II cohorts (Figure S1 and Figure S3).

In the HCP cognitive decoding task, IBSS-GAT consistently highlighted canonical large-scale functional systems known to support specific cognitive processes. Compared with the resting state, the default mode network (DMN) and frontoparietal control network (FPCN) showed significantly greater feature importance across multiple cognitive tasks, including relational reasoning, social cognition, working memory, and language processing (Figure S2). In addition, task-specific functional systems were selectively identified, including the limbic network (LMB) for emotion, the salience/ventral attention network (SVAN) for gambling and social cognition, the language network (LANG) for language processing, and the somatomotor network (SMN) for motor execution (see Supplementary Materials for details). These findings demonstrate that IBSS-GAT faithfully recapitulates well-established functional neuroanatomy underlying diverse cognitive processes.

Model interpretation further revealed biologically meaningful neural signatures with both convergent and disorder-specific characteristics. Across both ASD and MDD cohorts, IBSS-GAT consistently identified the DMN (ASD: Dice = 0.427, p_spin_ = 0.001; MDD: Dice = 0.427, p_spin_ = 0.001) and FPCN (ASD: Dice = 0.266, pspin = 0.006; MDD: Dice = 0.309, pspin = 0.001) as principal contributors to disease identification (Figure 2A, C and Tables S6–S7), whereas the DAN showed additional disease-specific importance for MDD (Dice = 0.244, p_spin_ = 0.002). Despite their shared involvement in disease identification at the group level, the two networks (FPCN and DMN) exhibited distinct associations with individual clinical phenotypes. In ASD, greater FPCN feature importance was significantly associated with overall symptom severity as well as social and communication impairments (ADOS Total: r = 0.215, p = 0.0003; Social: r = 0.210, p = 0.0004; Communication: r = 0.168, p = 0.005; Figure 2B), highlighting a preferential link between FPCN dysfunction and the core clinical manifestations of ASD. In contrast, in MDD, DMN feature importance significantly correlated with depression severity (HAMD Total: r = -0.110, p = 0.003, Figure 2D), whereas LMB feature importance was linked to anxiety severity (HAMA Total: r = 0.085, p = 0.022, Figure 2D), suggesting a predominant role of the DMN and LMB in affective dysregulation.

**Figure 2.**
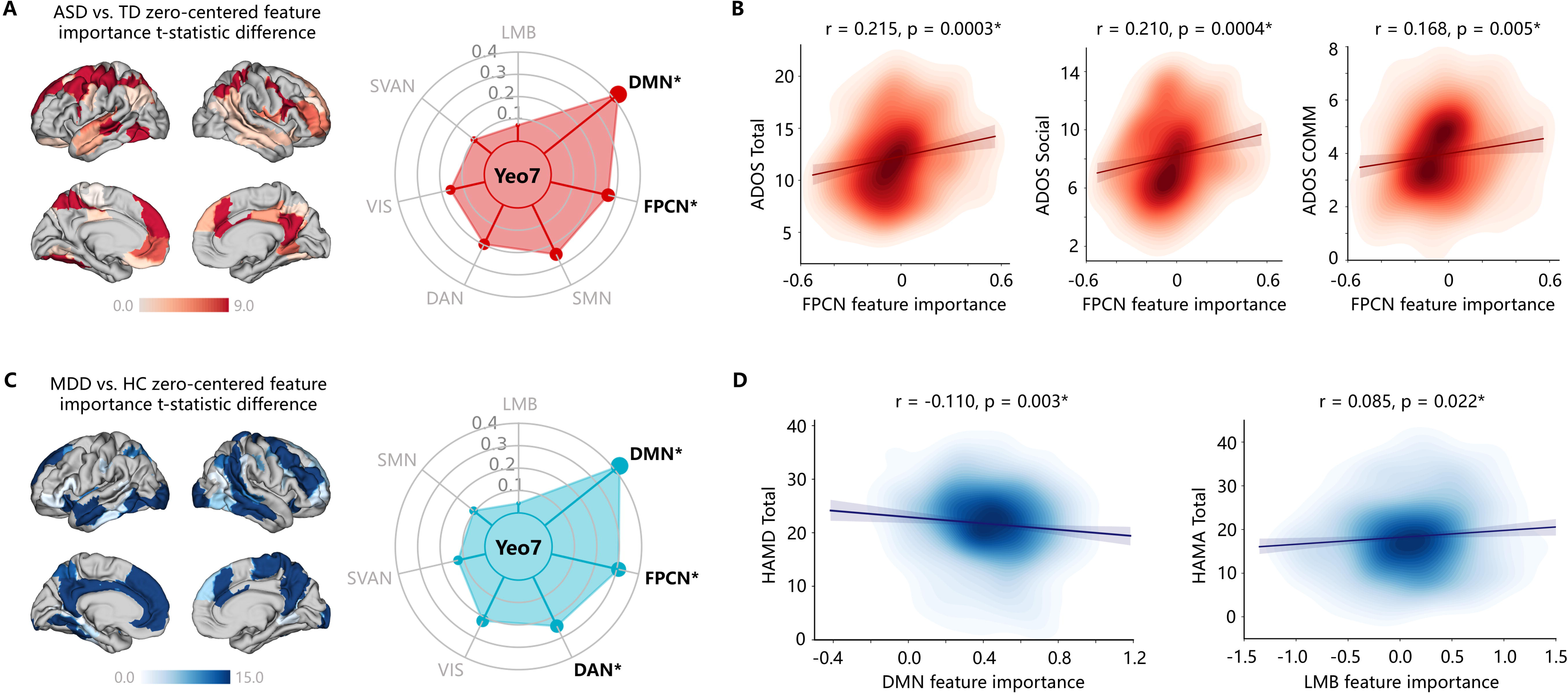
Model-derived predictive representations and their clinical associations in ASD and MDD. **(A)** In the ABIDE-I cohort, cortical maps show regions with greater GNNExplainer-derived, zero-centered feature importance in ASD than in TD, as indicated by positive t statistics. The radar plot summarizes their overlap with the Yeo seven-network parcellation. **(B)** Greater FPCN feature importance was associated with higher ADOS total, social, and communication scores in individuals with ASD. **(C)** Corresponding analysis in the DIRECT-II cohort showing regions with greater GNNExplainer-derived feature importance in MDD than in HC and their overlap with the Yeo seven-network parcellation. **(D)** DMN feature importance was negatively associated with HAMD total score, whereas limbic system (LMB) feature importance was positively associated with HAMA total score in individuals with MDD. Density contours illustrate the joint distribution of feature importance and clinical scores, and lines with shaded bands show fitted trends and 95% confidence intervals. Reported r and p values were obtained using Spearman correlation. Asterisks indicate significant network overlap or clinical associations. ASD, autism spectrum disorder; TD, typically developing; MDD, major depressive disorder; HC, healthy control; ADOS, Autism Diagnostic Observation Schedule; HAMD, Hamilton Depression Rating Scale; HAMA, Hamilton Anxiety Rating Scale; COMM, communication; DMN, default mode network; FPCN, frontoparietal control network.

### Metastate Reconstruction Bridges Individualized Neural Representations and Shared Disease Mechanisms

To interpret IBSS-derived individualized neural representations within a shared dynamic framework, we reconstructed the unified metastate space underlying individualized brain states across participants. Five metastates were consistently recovered in both the ABIDE-I and DIRECT-II datasets (Figure 3A). Despite being reconstructed independently from different cohorts and disorders, the resulting metastate spaces exhibited highly consistent spatial configurations, with significant correlations between corresponding metastate activation patterns ranging from 0.60 to 0.99 (Bonferroni-corrected p < 10^-10^). These findings suggest that individualized brain states can be organized within a unified metastate space, providing a shared dynamic reference for cross-subject mechanistic interpretation.

**Figure 3.**
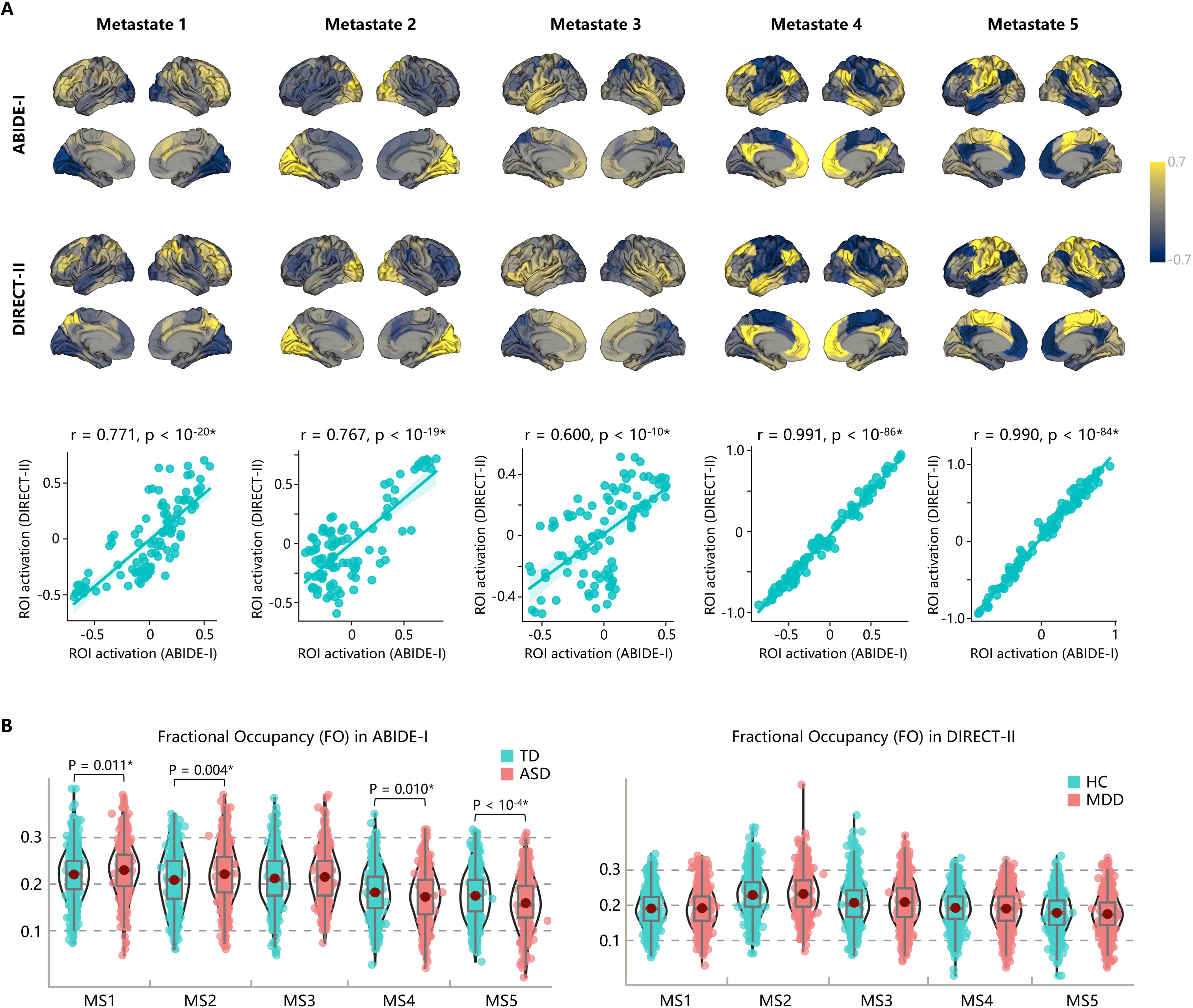
Cross-cohort correspondence of reconstructed metastate spaces and group differences in fractional occupancy. **(A)** Five metastates were independently reconstructed from individualized brain states in the ABIDE-I and DIRECT-II cohorts. Cortical maps show the activation patterns of corresponding metastates, and scatter plots compare their regional activation values across datasets. Pearson’s r and corresponding p values are shown above each plot. **(B)** Fractional occupancy of the five metastates in ASD and TD participants from ABIDE-I and in MDD and HC participants from DIRECT-II. Violin plots show the group distributions with individual participants overlaid. Significant group differences are annotated with p values. ROI, region of interest; FO, fractional occupancy; ASD, autism spectrum disorder; TD, typically developing; MDD, major depressive disorder; HC, healthy control.

Within this unified metastate space, ASD patients exhibited pronounced alterations in metastate dynamics. Specifically, ASD patients showed significantly higher fractional occupancy of Metastate 1 (T = 2.63, p = 0.011, FDR corrected) and Metastate 2 (T = 3.18, p = 0.004, FDR corrected), together with lower fractional occupancy of Metastate 4 (T = -2.76, p = 0.010, FDR corrected) and Metastate 5 (T = -4.57, p < 10^-4^, FDR corrected) than TD controls (Figure 3B). Among these altered metastate dynamics, the fractional occupancy of Metastate 1 emerged as a robust predictor of ADOS symptom severity (Total: r = 0.186, p = 0.001; Social: r = 0.181, p = 0.002; Figure 4A). Notably, this metastate significantly overlapped with the FPCN (Dice = 0.390, p_spin_ = 0.001) and SVAN (Dice = 0.302, p_spin_ = 0.001; Table S8), providing a dynamic interpretation for the prominent contribution of the FPCN identified by IBSS-GAT in ASD discrimination and phenotypic prediction.

**Figure 4.**
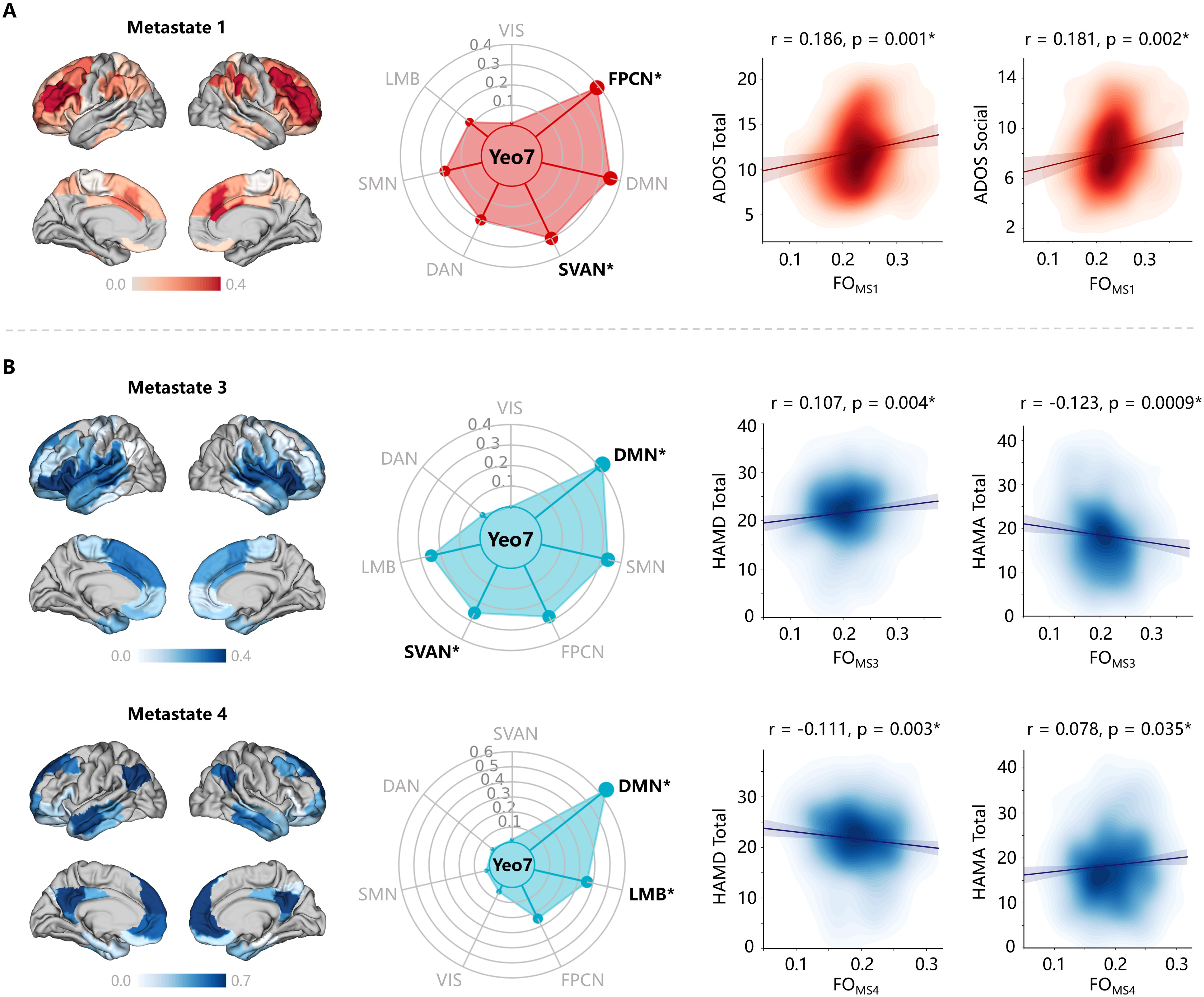
Metastate dynamics and their clinical associations in ASD and MDD. **(A)** In the ABIDE-I cohort, cortical maps show the activation pattern of Metastate 1, and the radar plot summarizes its overlap with the Yeo seven-network parcellation. Density plots show the associations between the fractional occupancy of Metastate 1 and clinical scores in individuals with ASD. **(B)** Corresponding analyses of Metastates 3 and 4 in the DIRECT-II cohort. Density contours illustrate the joint distributions of fractional occupancy and clinical scores; lines and shaded bands indicate fitted trends and 95% confidence intervals. Correlations were assessed using Spearman’s rank correlation. Asterisks indicate significant network overlap or clinical associations. FPCN, frontoparietal control network; SVAN, salience/ventral attention network; DMN, default mode network; LMB, limbic system; FO, fractional occupancy; ADOS, Autism Diagnostic Observation Schedule; HAMD, Hamilton Depression Rating Scale; HAMA, Hamilton Anxiety Rating Scale.

Although no significant group-level differences in metastate dynamics were observed between HC and MDD, these dynamics remained informative of individualized clinical phenotypes. The fractional occupancy of Metastate 3 predominantly involving the DMN (Dice = 0.421, p_spin_ = 0.038) and SVAN (Dice = 0.258, p_spin_ = 0.020; Table S9), was significantly associated with both HAMD (r = 0.107, p = 0.004) and HAMA (r = -0.123, p = 0.0009) scores (Figure 4B). In addition, the fractional occupancy of Metastate 4 was also linked to affective symptom severity (HAMD Total: r = -0.111, p = 0.003; HAMA Total: r = 0.078, p = 0.035) and exhibited joint activation of the DMN (Dice = 0.650, p_spin_ = 0.001) and LMB (Dice = 0.361, p_spin_ = 0.001; Table S10), indicating that MDD-related neural representations learned by IBSS-GAT are dynamically expressed as aberrant DMN–LMB dynamics.

### Independent Validation Confirms the Robustness and Reproducibility of IBSS framework

To evaluate the robustness and reproducibility of our framework, we independently replicated the complete analytical pipeline in the held-out multisite ABIDE-II cohort. Consistent with the findings in ABIDE-I, IBSS-GAT achieved comparable classification performance, with an ACC of 77.49 ± 2.44%, SEN of 74.41 ± 6.54%, SPE of 80.25 ± 6.91%, and AUC of 79.17 ± 4.02%, significantly outperforming conventional group-first metastate approaches and other state-of-the-art deep learning frameworks (Table 1).

Beyond predictive performance, the model-derived neural representations were also reproducible across independent cohorts (Figure S5A). GNNExplainer again identified prominent involvement of the DMN (Dice = 0.345, p_spin_ = 0.045) and FPCN (Dice = 0.262, p_spin_ = 0.022) in ASD discrimination (Table S11). Moreover, greater feature importance of the FPCN was significantly associated with more severe autistic symptoms (ADOS-2 Total: r = 0.172, p = 0.040), reproducing the individualized neural signatures observed in ABIDE-I.

We further examined whether the dynamic interpretations of these neural representations were reproducible across datasets (Figure S5B). Reconstruction of the unified metastate space from the ABIDE-II IBSSs again yielded 5 metastates. Among them, the fractional occupancy of an FPCN-dominated metastate (Dice = 0.349, p_spin_ = 0.005, Table S12) was positively associated with ADOS-2 total scores (r = 0.194, p = 0.020), providing independent support for the reproducibility of symptom-related FPCN metastate dynamics in ASD.

Taken together, these findings demonstrate that IBSS achieves reproducible predictive performance, individualized neural representations, and dynamic mechanistic interpretations across independent cohorts, supporting the robustness and generalizability of the proposed framework.

## DISCUSSION

A defining challenge in computational psychiatry is reconciling two competing goals: preserving inter-individual neurobiological heterogeneity for personalized prediction, and identifying generalizable, phenotype-associated neural features for mechanistic insight. In this study, we developed IBSS as an individual-first framework that enables individualized brain state modeling while allowing shared dynamic patterns to be recovered across participants. By first preserving individual-specific brain state configurations, IBSS-GAT achieved state-of-the-art performance in cognitive decoding and psychiatric disorder identification across four large-scale neuroimaging cohorts, with accuracy improvements of 16.66% to 36.69% over conventional group-first metastate approaches. Subsequent recovery of the unified metastate space further organized these individualized states within a common dynamic reference and translated model-derived representations into interpretable dynamic phenotypes, revealing reproducible hyper-occupancy of FPCN in ASD and symptom-related DMN–limbic dynamics in MDD. These findings demonstrate that individualized state estimation and shared dynamic interpretation are not mutually exclusive. By separating them into sequential stages, IBSS links individual-level prediction to cross-subject neurodynamic mechanisms, providing a general framework for dissecting heterogeneity in psychiatric disorders.

The primary advantage of IBSS-GAT arises from the individualized brain-state representation rather than from model complexity alone. Conventional metastate approaches estimate a single shared state repertoire across participants, compressing inter-individual variability in spatial state organization into group-level centroids and limiting individual differences primarily to temporal metrics such as fractional occupancy, dwell time, and transition probability (10, 12–24, 35, 36). The comparison with MS-GAT is therefore particularly illuminating: holding the graph attention architecture constant, replacing IBSS with group-level metastates reduced accuracy by 16.66–36.69 percentage points across all cohorts (Table 1). Ablation analyses further confirmed that individualized activation patterns—including idiosyncratic spatial configurations and adaptively determined state counts—were the dominant source of discriminative information, while state transition topology and attention-based aggregation provided complementary but smaller contributions (Table 2). This hierarchy indicates that the limited predictive utility of conventional dynamic metrics arises partly from information lost during group-level state estimation, which cannot be fully recovered by quantifying temporal dynamics around shared centroids.

Critically, cross-subject comparability can be recovered from individualized state representations after their independent estimation. The strong spatial correspondence between metastate spaces independently reconstructed in ABIDE-I and DIRECT-II supports the broader rationale for bottom-up shared reconstruction (Figure 3A). Despite differences in cohort composition and diagnosis, corresponding metastates showed highly consistent activation patterns (correlations ranging from 0.60 to 0.99), suggesting that heterogeneous individualized states can converge onto a reproducible lower-dimensional organization. This is consistent with accumulating evidence that functional brain organization contains robust individual-specific features while retaining shared large-scale organizational principles (25, 37). Importantly, this correspondence emerged only after the participant-specific state spaces had been estimated, rather than through the prior imposition of a common state repertoire (10, 12, 25). The finding therefore indicates that cross-subject comparability can be recovered without constraining the initial individual-level representation. This does not imply a fixed, universal metastate library (25, 38, 39). The reconstructed organization may vary with preprocessing, parcellation, state estimation, and clustering choices (39, 40). Instead, its significance lies in demonstrating that a comparable dynamic reference can emerge from independently estimated individual states. The reconstructed unified metastate space thus functions as a data-derived reference coordinate system: it preserves the individualized representations used for prediction while enabling systematic cross-subject comparison and dynamic mechanistic interpretation.

The combination of feature attribution and metastate reconstruction further addresses a longstanding limitation of deep learning in neuroimaging: the gap between high predictive performance and biologically meaningful interpretation (41–44). To address this, we adopted a two-stage interpretive strategy: feature attribution identified the regional and network patterns that contributed to IBSS-GAT predictions, whereas metastate reconstruction showed how related functional configurations were dynamically expressed through fractional occupancy. These analyses therefore provide complementary views of the same individualized representations: one locates the neural features used by the model, and the other places those features within an interpretable temporal organization. The HCP-task results offered a useful positive control for this interpretive framework. IBSS-GAT recovered canonical systems associated with higher-order cognition (45–47), emotion (48), language (49), gambling (50), and motor execution (51), indicating that the learned representations retained recognizable functional neuroanatomy rather than merely supporting statistical discrimination (see the Supplementary Materials for details). In clinical cohorts, the convergence between network feature importance, metastate occupancy, and symptom severity further grounded predictive signatures in well-characterized neurobiological processes.

In ASD, converging evidence from model interpretation and metastate dynamics points to a reproducible neurodynamic signature centered on FPCN hyper-engagement, expressed dynamically through increased occupancy of an FPCN–SVAN coactivation state. Given the roles of the FPCN in flexible coordination across distributed systems (46, 52) and of the SVAN in detecting behaviorally relevant information (53, 54), their excessive co-engagement may reflect atypical control–salience coordination within the broader triple-network system (55, 56), potentially disrupting the balance between externally directed control and DMN-related internal processing in ASD (57). Importantly, this signature was replicated across independent cohorts at the predictive, representational, dynamic, and clinical levels, reducing the likelihood of cohort-specific artifacts and supporting the robustness of FPCN-related neurodynamic alterations in ASD.

In MDD, the findings revealed a distinct DMN-LMB pattern in which clinically informative neural dynamics manifested at the dimensional symptom level rather than as a uniform diagnostic group effect. Given the roles of the DMN in self-referential and internally oriented cognition (58, 59) and of the limbic network in affective valuation and emotional reactivity (60–62), their co-activation may indicate altered integration between internally generated thought and affective valuation in MDD, potentially biasing self-focused processing toward mood-congruent content and contributing to individual variation in depressive and anxiety symptoms (63–65). Notably, these symptom-linked dynamics were observed despite the absence of significant group-level differences in metastate occupancy, highlighting that diagnostically defined categories can mask continuous neurobiological variation (66, 67). This reinforces the rationale for individual-first representations: clinically meaningful neural information may exist at the individual and dimensional level even when it does not emerge as a uniform patient–control contrast (68).

Several limitations should be acknowledged. First, individual-level HMMs estimated from relatively short time series may yield uncertainty that varies across participants, particularly when the inferred number of states is large relative to the available observations. Although the present framework produced robust results across datasets and parcellation granularities, future work could incorporate hierarchical or uncertainty-aware models to stabilize individual state estimation while preserving idiosyncrasy, and test–retest analyses to dissociate trait-like individual features from transient state fluctuations. Second, the ABIDE-II analysis independently replicated the complete pipeline and the main ASD findings, but it did not constitute external validation of a locked model because the classifier was retrained within that cohort. Leave-one-site-out evaluation, direct cross-cohort model transfer, and prospective validation using harmonized protocols will be needed to establish generalizability to unseen settings. Third, feature attribution results may be influenced by correlated neural features and model architecture, and neither feature importance nor symptom association establishes causal neural mechanisms. Longitudinal, interventional, and perturbation-based studies will be required to test causal and prognostic relevance. Fourth, the current metastate reconstruction relied primarily on mean activation patterns; incorporating covariance structure, weighted transition probabilities, and higher-order temporal context may reveal additional shared organization (22). Finally, the analyses were cross-sectional, the brain–symptom associations were modest, and the MDD findings lacked independent replication. Whether IBSS-derived representations can predict treatment response, relapse, disease progression, or longitudinal symptom change remains to be determined.

In conclusion, the present study proposed an individual-first brain state modeling framework that reconciles individualized prediction with generalizable neurodynamic interpretation in psychiatric disorders. By jointly modeling the spatiotemporal architecture of individual-specific brain state spaces, our model consistently outperformed existing deep learning frameworks and achieved substantial gains over conventional group-first metastate approaches. Reconstruction of the unified metastate space further linked individualized predictive representations to biologically interpretable metastate dynamics, delineating both transdiagnostic commonalities and clinically distinct signatures across psychiatric disorders. More broadly, this individual-first, group-reconstruction strategy provides a flexible, generalizable framework for characterizing psychiatric heterogeneity and may support future applications in dimensional phenotyping and clinical stratification.

## METHODS

### Study Cohorts

#### HCP-task Cohort

We used the HCP-task dataset to validate the representation and classification capabilities of IBSS-GAT. The fMRI scans in this dataset are collected from 590 healthy adults as part of the Human Connectome Project (69). For each subject, recordings are taken during seven different cognitive tasks (emotion, gambling, language, motor, relational, social and working memory) and also during rest (see Table S1 and (70) for details of task designs).

#### ABIDE Cohorts

Resting-state fMRIs and phenotypic information from the multisite ABIDE databases (including ABIDE-I (71) and ABIDE-II (72)) were utilized in this study. All fMRI data underwent preprocessing and quality control. The resulting sample comprised 436 autism spectrum disorder (ASD) patients and 470 typically developing (TD) controls from ABIDE-I; and 324 ASD patients, 369 TD controls from ABIDE-II. Participant demographic information is summarized in Table S2. Analyses were conducted independently on ABIDE-I and ABIDE-II to verify the robustness of our framework and the reproducibility of the findings.

#### DIRECT-II Cohort

This database comprises resting-state fMRI scans of 1660 patients with major depressive disorder (MDD) and 1341 healthy controls (HCs) form 23 different sites (https://rfmri.org/DIRECT-Phase2) (73, 74). All participants are East Asian, and patients’ diagnoses are based on the International Classification of Diseases, 10th revision (ICD-10) or Diagnostic and Statistical Manual of Mental Disorders, 4th edition (DSM-IV) criteria. Our analyses were restricted to 1283 MDDs and 1118 HCs who had a sufficient number of usable fMRI frames. Participant demographic information is summarized in Table S3.

### fMRI Data Preprocessing

Imaging data form HCP-task were preprocessed following (75). Specifically, brain data were normalized to fslr32k space using MSM-AII registration and parcellated into 100 regions (76). Nuisance signals were removed using CompCor (five principal components from white matter and ventricular masks), along with 12 detrended motion parameters provided by the Human Connectome Project. The global signal was regressed out, and time series were band-pass filtered (0.009–0.08 Hz). Frames with framewise displacement >0.2 mm or DVARS >75 were censored, and sessions with more than 50% censored frames were excluded. For each subject and scan, analyses were restricted to the first 176 time points (corresponding to the shortest scan of the emotion task) to control for differences in task duration.

All imaging data from ABIDE cohorts were preprocessed using *fMRIPrep* pipeline (v25.1.1; https://fmriprep.org/en/stable/) (77). In brief, functional data were processed by slice timing correction using *3dTshift* from *AFNI* (78), motion correction using *FSL-mcflirt* (79), fieldmap-less susceptibility distortion correction, BOLD-to-T1w transformation using *bbregister* in *FreeSurfer* with nine degrees of freedom (80), and resampling to 2mm isotropic MNI space. These preliminary processing stages were then followed by the post-processing and noise regression pipeline *XCP-D* (v0.11.0; https://xcp-d.readthedocs.io/en/latest/) (81). Post-processing steps included nuisance covariates regression (including 6 motion parameters, mean global signal, mean WM signal, mean CSF signal with their temporal derivatives, and quadratic expansion of 6 motion parameters, tissue signals, and their temporal derivatives), high-motion volumes censoring (FD > 0.5 mm or DVARS > 1.5 SD above mean), temporal interpolation (cubic spline), bandpass filtering (0.01-0.1Hz), and spatial smoothing (full width at half maximum = 6mm). Participants lacking sufficient usable time points (<150 TR) were excluded. Regional averaged functional timeseries were extracted from the processed BOLD signals using the Schaefer atlas (76) with 100 regions of interest (ROIs). For each subject, we only analyzed the first 150 measurements in timeseries to control data size variability.

The resting-state fMRI data from the DIRECT-II Cohort had undergone standardized preprocessing using the DPABISurf pipeline (82–84) prior to public release. The preprocessing procedures included removal of the initial volumes, slice-timing correction, head-motion correction, spatial normalization to standard space, nuisance regression, surface projection, and quality-control procedures, as detailed in the DIRECT Phase II data-sharing protocol (74). Regional mean time series were extracted according to the Schaefer atlas (76) with 100 ROIs. As an additional preprocessing step, we performed global signal regression (GSR) on the regional time series to further reduce global non-neuronal fluctuations. To minimize variability in time-series length across subjects, analyses were restricted to the first 150 time points of each scan, and scans with less than 150 slices were filtered out, leaving 1283 MDDs and 1118 HCs.

### Construction of the Individual-specific Brain State Space (IBSS)

Conventional brain state identification approaches typically derive a common set of metastates across subjects through group-level modeling, obscuring substantial inter-individual variability in brain state organization. To better capture subject-specific functional architectures for individualized prediction, we developed an Individual-Specific Brain State Space (IBSS) framework that explicitly models individualized brain-state representations.

To construct the IBSS, we first applied a hidden Markov model (HMM) on each participant to identify individual-specific brain states, where each state corresponds to a unique pattern of whole-brain spontaneous activity, modeled by a multivariate Gaussian distribution and a state time course indicating the points in time at which that state is active (12, 85, 86). Specifically, if vector ***x_t_*** represents the observed BOLD signal and q_t_ represents the hidden state at time point t, the observational model is assumed as:

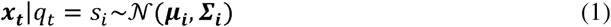

where ***µ_i_*** is an N-dimensional vector containing the mean BOLD activation across *N* ROIs and ***Σ_i_*** denotes the corresponding diagonal covariance matrix describing regional variability when state *s_i_* is active. Furthermore, the state sequence is constrained by modeling the transition probabilities between all pairs of brain states, such that the probability of a given state at time point t depends on the state that is active at time point *t*- 1:

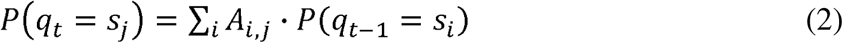

where *A_i,j_* refers to the transition probability from state *s_i_* to state *s_j_*. Given a parameter ***π*** encoding the initial state probabilities for each scanning session, the observed data at each time point is effectively modeled as a mixture of Gaussian distributions weighted by *w_t_* (*_i_*) = *P*(*q_t =_ s_i_*).

Unlike most studies applying HMM to all concatenated data (group-level) (12, 17, 35, 36, 87), we inferred HMM on each participant independently to highlight individual differences in state space. For each sample, we repeated the HMM modeling process with varying *k* (number of states) between 3 and 60, and selected the optimal number of states as the *k* yielding the minimum Akaike information criterion (AIC), defined as

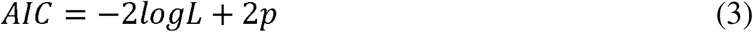

where *L* denotes the maximized likelihood of the observed data and *p* represents the number of free model parameters (88, 89). By balancing goodness of fit against model complexity, this criterion retains sufficient model flexibility to capture subject-specific state-space complexity while reducing the risk of overfitting. The HMMs were inferred using the *hmmlearn* package, which provides estimates of the parameters of the state distribution *N*(*µ*,*E*), the transition probability matrix and the hidden state time course (see the Supplementary Materials for details).

To fully capture the spatiotemporal structure of this latent dynamics, the IBSS was formulated as a directed graph:

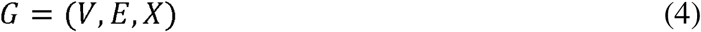

where *V* denotes the set of latent brain states identified by the individual HMM, *E* denotes the transition relationships among these states, and *x* represents the node feature matrix. For a participant with *k* latent states, the node set if defined as:

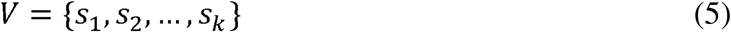

Each node *s_i_* corresponds to a latent brain state and is associated with an activation pattern

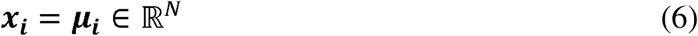

where *N* is the number of ROIs and *µ_i_* is the mean BOLD activation vector estimated by the HMM. The node feature matrix is therefore given by

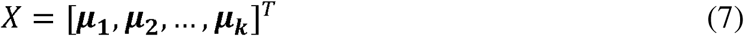

The graph edges were derived from the individual-specific state transition matrix *A*. Specifically, a directed edge from state *s_i_* to state *s_j_* was assigned whenever a transition between the two states was observed in the hidden-state time course:

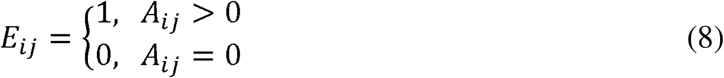

This binarization yielded a directed adjacency matrix that captures the topology of reachable state transitions. Consequently, the IBSS represents each participant as a unique graph structure that jointly encodes the spatial activation patterns of latent brain states and their temporal transition dynamics, providing a compact characterization of individualized state-space organization (Figure 1B). These participant-specific IBSS graphs were constructed using the *PyTorch Geometric* package.

### IBSS-GAT for Cognitive Decoding and Disease Identification

To evaluate the predictive utility of the individual-specific brain state space, we assessed its performance across both cognitive and clinical prediction tasks. In particular, we applied IBSS to four independent datasets (HCP-task, ABIDE-I, ABIDE-II and DIRECT-II cohorts) and examined its ability to support cognitive decoding as well as disease identification, including ASD and MDD.

To leverage the IBSS for predictive modeling, we employed a 2-layer Graph Attention Network (GAT) (90) using the *PyTorch* toolkit, with two attention heads in the first layer and a single attention head in the second layer. The hidden feature dimension was set to 64 for the HCP-task cohort and 32 for the ABIDE-I, ABIDE-II, and DIRECT-II cohorts. For each participant-specific IBSS graph, the GAT computed node representations by adaptively aggregating features from directly connected neighboring states. Specifically, each node aggregated features from its neighboring states, with the corresponding attention coefficients learned through a shared self-attention mechanism. Rectified linear unit (ReLU) activation was applied after each graph attention layer, and dropout with a rate of 0.5 was applied to the normalized attention coefficients to reduce overfitting. The resulting node embeddings were then averaged across the graph using global mean pooling to generate an individual-level feature vector, which was subsequently passed to a fully connected linear classifier to produce task-specific predictions (see the Supplementary Materials for details).

For the HCP-task cohort, models were trained to classify 7 cognitive tasks and the resting state; for the ABIDE-I and ABIDE-II cohorts, to distinguish individuals with ASD from TD controls; and for the DIRECT-II cohort, to distinguish individuals with MDD from HCs. Accordingly, the output dimension of the linear classifier was set to eight for the HCP-task cohort and two for the clinical cohorts. Model parameters were optimized using the Adam optimizer with a learning rate of 0.01, and cross-entropy loss was used for all classification tasks. Batch sizes were set to 64 for HCP-task and 32 for ABIDE-I, ABIDE-II and DIRECT-II. For evaluation, we employed a 10-fold cross-validation strategy, in which participants were randomly partitioned into 10 non-overlapping folds, with each fold used once as the test set. For the HCP-task cohort, splitting was performed at the participant level, such that all task and resting-state samples from the same participant were assigned to the same fold. Classification performance was measured by accuracy (ACC), sensitivity (SEN), specificity (SPE) and area under receiver operating characteristic curve (AUC).

To quantify the benefits of enhanced state individualization enabled by IBSS, we compared its predictive performance with that of conventional group-first metastate approaches. Specifically, HMM was applied to the concatenated BOLD time series from all subjects to identify a common set of metastates shared across individuals, with the number of metastates matched to that used in the IBSS framework. Each subject was also represented as a state transition graph using these shared state activation patterns as graph nodes, such that inter-individual differences were encoded solely by transition dynamics (edges). To ensure a fair comparison, the same two-layer GAT architecture was applied to these metastate representations across all prediction tasks, yielding the MS-GAT model. Besides, based on the group-level metastates, we also calculated conventional brain-state metrics, including state fractional occupancy (FO), dwell time (DT) and transition probabilities (TP), which served as input features of a support vector machine (SVM) for classification and comparison with the proposed IBSS-GAT. In addition, the proposed IBSS-GAT was also benchmarked against several state-of-the-art deep learning models, including BrainGNN (91), ST-GCN (92), dFCExpert (93) and Brain Network Transformer (94).

### Ablation Studies

We conducted ablation experiments to assess the contributions of individual components of IBSS-GAT, specifically the activation patterns and transition patterns in the IBSS, and the GAT. For the ablation of activation patterns, we generated the same number of random activation patterns for each subject to replace the original state activation patterns as graph nodes, while preserving the original state transition dynamics (edges). To ablate the transition patterns, the original state transition matrix of each subject was replaced with a randomly generated binary matrix matched in sparsity (graph edges), while keeping the original state activation patterns (graph nodes) unchanged. Finally, for GAT ablation, we used a 2-layer graph convolution network (GCN) (95) instead of GAT to learn graph-level representations from the IBSS.

### Interpretation of Cognitive Representations Learned by IBSS-GAT

We used a model-agnostic GNN explainability method—GNNExplainer (34)—to measure the feature importance associated with task classification in each brain region and for each sample. The derived feature attribution maps of each task (Figure S1) were compared with that of rest using pairwise t-tests, and the spatial profile of brain regions with task-rest t-statistic difference (T value) >0 was mapped to canonical functional networks (17 instrinsic functional connectivity networks according to Yeo et al. (96) and 12 Cole-Anticevic networks (CAB-NP) defined by Ji (97), which complement each other, collectively covering all major networks implicated in cognitive processing) using the Network Correspondence Toolbox (NCT) (98) to identify subnetworks that discriminated between rest and each cognitive task. Specifically, for each functional network, spatial correspondence with the task-rest difference map was assessed as the Dice coefficient of network overlap, and spin tests with 1,000 permutations was used to quantify levels of significance (98).

### Interpretation of Disorder Representations Learned by IBSS-GAT

We also used the GNNExplainer (34) to quantify the feature importance of individual brain regions for ASD versus TD classification. A t-statistic map was generated by comparing feature importance between ASD and TD participants. Regions exhibiting greater importance in ASD than TD (T value > 0) were mapped onto the seven intrinsic functional connectivity networks defined by Yeo et al. (96) using the NCT (98) to identify functional networks that discriminated between the ASD and TD groups. Significant network overlap was set at p_spin_ < 0.05 (1,000 permutations). Furthermore, we explored the relationship between the model-identified network features and the severity of clinical symptoms in ASDs. Spearman correlations between the ADOS scores and the average feature importance of each subnetwork were computed. The significance threshold was set at p < 0.05 with false discovery rate (FDR) correction.

The same interpretability pipeline was applied to the MDD-HC classification task. Clinical relevance was assessed by examining the associations between network-level feature importance and symptom severity as measured by the Hamilton Depression Rating Scale (HAMD) and Hamilton Anxiety Rating Scale (HAMA).

### Reconstruction of the Unified Metastate Space Underlying IBSS

Unlike conventional metastate identification methods that directly cluster fMRI time points across all subjects, our framework recovers the underlying cross-subject metastate space by applying k-means clustering to individualized brain states from all participants’ IBSSs in ABIDE-I, grouping states with similar activation patterns into shared metastates. This bottom-up reconstruction strategy preserves individualized neural representations while organizing them within a unified dynamic framework. The optimal number of metastates was selected via the Krzanowski-Lai criterion (supplemented by the Davies-Bouldin Index (99) and the Silhouette score (100); Figure S4) (101), given its effectiveness in determining topographic map partitions based on quality metrics and overall explained variance (102–104). Fractional occupancy, which denotes the total proportion of time a system spends in a specific metastate or the probability of being in that metastate, was measured and compared between ASD and TD groups using two-sample t-tests (normally distributed data) and Mann-Whitney U tests (non-normally distributed data). Relationships between the metastate fractional occupancy and clinical symptoms in ASD were assessed using Spearman’s correlations. Significance of all statistical analyses was set at FDR corrected p < 0.05.

The same procedure for reconstructing metastate space, characterizing metastate dynamics, and assessing their clinical relevance was independently performed on the DIRECT-II dataset. Furthermore, to evaluate whether independently reconstructed metastate spaces converged to a unified metastate space across datasets and disorders, corresponding metastates derived from the ABIDE-I and DIRECT-II datasets were compared by calculating Pearson correlation coefficients between their activation patterns. Statistical significance was determined at Bonferroni-corrected p < 0.05.

### Sensitivity and Validation Analyses

We performed both sensitivity and validation analyses to examine the robustness of our method and results. First, IBSS-based classification performance was revaluated based on different atlases across a range of parcellation granularities (Schaefer atlas with 200, 400 parcels (76)). Second, data from ABIDE-II were used to validate the reproducibility and reliability of the ASD-related findings revealed by our framework. Specifically, we independently repeated the entire analytical pipeline on the ABIDE-II cohort, including IBSS-based classification, model interpretation, and metastate dynamic analyses.

## Data Availability

All data produced are available online at: https://www.humanconnectome.org/study/hcp-young-adult, https://fcon_1000.projects.nitrc.org/indi/abide/, and https://rfmri.org/DIRECT-Phase2

## Data availability

Data from the HCP, ABIDE-I, ABIDE-II, and DIRECT-II databases are made available via: https://www.humanconnectome.org/study/hcp-young-adult, https://fcon_1000.projects.nitrc.org/indi/abide/, and https://rfmri.org/DIRECT-Phase2.

## Code availability

Code for the IBSS framework will be released in https://github.com/Kecici/IBSS. Code for other statistical analyses and visualizations will be made available from the corresponding author, upon reasonable request.

## Acknowledgements

This study is supported by grants from the National Key Research and Development Program of P.R. China (2025YFF0517803), the National Natural Science Foundation of P.R. China (62276081), Guangdong S&T Programme (2025B0101130004), and the Shenzhen Science and Technology Program (GXWD20231129121139001, JCYJ20240813110522029, CJGJZD20230724093959002, JCYJ20250604145427037).

We greatly appreciate the contributions of the Autism Brain Imaging Data Exchange, the Depression Imaging REsearch ConsorTium, and the Human Connectome Project without which this work would not be possible.

## Author Contributions

T.M. supervised the whole work. C.Y. and T.M. revised the manuscript and financially supported the study. C.R. developed the analytical methods, prepared the figures, and drafted the manuscript. C.R. and X.L. analyzed the data.

## Competing interests

The authors declare no competing interests.

## Declaration of generative AI in the writing process

During the preparation of this work, ChatGPT-5.6 was used solely for grammar correction and language polishing. No core academic content was generated by the tool. All AI-assisted content was fully reviewed and edited by the authors, who take full responsibility for the entire content of this publication.

## Notes

### Competing Interest Statement

The authors have declared no competing interest.

### Author Declarations

The study used (or will use) ONLY openly available human data that were originally located at: https://www.humanconnectome.org/study/hcp-young-adult, https://fcon_1000.projects.nitrc.org/indi/abide/, and https://rfmri.org/DIRECT-Phase2

### Summary of Updates

We have substantially reconceptualized and expanded the manuscript. Specifically, we have: 1) reframed IBSS as an individual-first, group-reconstruction framework addressing the trade-off between personalized prediction and shared neurodynamic interpretation; 2) extended the study from ASD to both ASD and MDD with additional large-scale validation, matched baseline comparisons, and cross-disorder metastate reconstruction; and 3) clarified the model development and evaluation procedures, including architecture, optimization, participant-level 10-fold cross-validation, and benchmark settings; 4) all figures revised and Supplemental files updated.

