## Supplementary Materials for "Individual-First Brain State Modeling Reconciles Personalized Prediction with Generalizable Mechanistic Interpretation in Precision Psychiatry"

### Supplementary material

**HMM parameter estimation: IBSS activation and transition patterns**

The state distribution $\mathcal{N}(\boldsymbol{\mu}_{\boldsymbol{i}},\boldsymbol{\Sigma}_{\boldsymbol{i}})$, transition probability matrix $\boldsymbol{A}$, and initial state probabilities $\boldsymbol{\pi}$ are estimated by the forward-backward algorithm (1, 2), which defines a forward probability (the joint probability of observing the first $t$ observations and being in state $s_{j}$ at time point $t$) as:

$\alpha_{t}\left( j \right)=P\left( \boldsymbol{x}_{\boldsymbol{1}},\boldsymbol{x}_{\boldsymbol{2}},...,\boldsymbol{x}_{\boldsymbol{t}},q_{t}=s_{j} | \boldsymbol{\theta} \right)=\left\{ \begin{aligned} \pi_{j}\cdot\mathcal{N}\left( \boldsymbol{x}_{\boldsymbol{t}} | \boldsymbol{\mu}_{\boldsymbol{j}},\boldsymbol{\Sigma}_{\boldsymbol{j}} \right) t=1 \\ \mathcal{N}\left( \boldsymbol{x}_{\boldsymbol{t}} | \boldsymbol{\mu}_{\boldsymbol{j}},\boldsymbol{\Sigma}_{\boldsymbol{j}} \right)\sum_{i=1}^{k} \alpha_{t-1}\left( i \right)A_{i,j} t=2,3,\ldots,T \end{aligned} \right.$(1)

and a backward probability (Given being in state $s_{i}$ at time point $t$, the conditional probability of observing from $t+1$ to the end) as:

$\beta_{t}\left( i \right)=P\left( \boldsymbol{x}_{\boldsymbol{t+1}},\boldsymbol{x}_{\boldsymbol{t+2}},...,\boldsymbol{x}_{\boldsymbol{T}} | q_{t}=s_{i},\boldsymbol{\theta} \right)=\left\{ \begin{aligned} 1 t=T \\ \sum_{j=1}^{k} {\mathcal{N}(\boldsymbol{x}_{\boldsymbol{t+1}}|\boldsymbol{\mu}_{\boldsymbol{j}},\boldsymbol{\Sigma}_{\boldsymbol{j}})\beta}_{t+1}(j)A_{i,j} t=1,2,\ldots,T-1 \end{aligned} \right.$(2)

where $\boldsymbol{X}=\boldsymbol{[x}_{\boldsymbol{1}},\boldsymbol{x}_{\boldsymbol{2}},...,\boldsymbol{x}_{\boldsymbol{T}}]$ is the observed signal, $\boldsymbol{\theta}=(\boldsymbol{A},\mathcal{N}(\boldsymbol{\mu},\boldsymbol{\Sigma}),\boldsymbol{\pi})$ are the current model parameters, $k$ is the number of hidden states, thus the probability $\gamma_{t}(i)$ of being in state $s_{i}$ and the joint probability $\xi_{t}(i,j)$ of transition from state $s_{i}$ to state $s_{j}$ at time point $t$ can be calculated as:

$\gamma_{t}\left( i \right)=P\left( q_{t}=s_{i} | \boldsymbol{X},\boldsymbol{\theta} \right)=\frac{\alpha_{t}\left( i \right)\beta_{t}\left( i \right)}{\sum_{j=1}^{k} \alpha_{t}\left( j \right)\beta_{t}\left( j \right)}$(3)

$\xi_{t}(i,j)=P(q_{t}=s_{i},q_{t+1}=s_{j}|\boldsymbol{X},\boldsymbol{\theta})=\frac{\alpha_{t}(i)A_{i,j}{\mathcal{N}(\boldsymbol{x}_{\boldsymbol{t+1}}|\boldsymbol{\mu}_{\boldsymbol{j}},\boldsymbol{\Sigma}_{\boldsymbol{j}})\beta}_{t+1}(j)}{\sum_{i,j} \alpha_{t}(i)A_{i,j}{\mathcal{N}(\boldsymbol{x}_{\boldsymbol{t+1}}|\boldsymbol{\mu}_{\boldsymbol{j}},\boldsymbol{\Sigma}_{\boldsymbol{j}})\beta}_{t+1}(j)}$(4)

Parameters $\boldsymbol{\mu}_{\boldsymbol{i}}$, $\boldsymbol{\Sigma}_{\boldsymbol{i}}$ and $A_{i,j}$ are updated according to $\gamma_{t}(i)$ and $\xi_{t}(i,j)$:

$\boldsymbol{\mu}_{\boldsymbol{i}}^{\boldsymbol{new}}=\frac{\sum_{t=1}^{T} \gamma_{t}(i)\boldsymbol{x}_{\boldsymbol{t}}}{\sum_{t=1}^{T} \gamma_{t}(i)}$(5)

$\boldsymbol{\Sigma}_{\boldsymbol{i}}^{\boldsymbol{new}}=\frac{\sum_{t=1}^{T} \gamma_{t}(i)(\boldsymbol{x}_{\boldsymbol{t}}-\boldsymbol{\mu}_{\boldsymbol{i}}^{\boldsymbol{new}}){(\boldsymbol{x}_{\boldsymbol{t}}-\boldsymbol{\mu}_{\boldsymbol{i}}^{\boldsymbol{new}})}^{T}}{\sum_{t=1}^{T} \gamma_{t}(i)}$(6)

$A_{i,j}^{new}=\frac{\sum_{t=1}^{T-1} \xi_{t}(i,j)}{\sum_{t=1}^{T-1} \gamma_{t}(i)}$(7)

until the change in log-likelihood $\log P(\boldsymbol{X}\left| \boldsymbol{\theta} \right.)$ is less than a threshold, at which point the HMM model achieves optimal fit and the $k$ hidden states characterized by activation patterns $\boldsymbol{\mu}_{\boldsymbol{1}}^{\boldsymbol{*}},\boldsymbol{\mu}_{\boldsymbol{2}}^{\boldsymbol{*}},...,\boldsymbol{\mu}_{\boldsymbol{k}}^{\boldsymbol{*}}$ are identified.

The hidden state time series $\boldsymbol{q}^{\boldsymbol{*}}=\left[ q_{1}^{*},q_{2}^{*},...,q_{T}^{*} \right]$ are decoded using the Viterbi algorithm (3), which seeks the global optimal path $\boldsymbol{q}^{\mathbf{*}}$ maximizing the joint probability $P(\boldsymbol{X},\boldsymbol{q}|\boldsymbol{\theta}^{\mathbf{*}})$ through dynamic programming. We recursively calculate the maximum joint probability among all paths that can generate the first $t$ observations ending in state $s_{j}$：

$\left\{ \begin{matrix} \delta_{t}(j)=\pi_{j}\mathcal{\cdot N(}\boldsymbol{x}_{\boldsymbol{t}}|\boldsymbol{\mu}_{\boldsymbol{j}},\boldsymbol{\Sigma}_{\boldsymbol{j}}) t=1 \\ \delta_{t}(j)=\max_{1\leq i\leq k} [\delta_{t-1}(i)\cdot A_{i,j}\mathcal{]\cdot N(}\boldsymbol{x}_{\boldsymbol{t}}|\boldsymbol{\mu}_{\boldsymbol{j}},\boldsymbol{\Sigma}_{\boldsymbol{j}}) t=2,3,...,T \end{matrix} \right.$	(8)

then a path pointer instructing the previous state at $t-1$ that maximize $\delta_{t}(j)$ is defined:

$\left\{ \begin{matrix} \psi_{t}(j)=0 t=1 \\ \psi_{t}(j)=\underset{1\leq i\leq k}{\mathrm{argmax}} [\delta_{t-1}(i)\cdot A_{i,j}] t=2,3,...,T \end{matrix} \right.$	(9)

thus the optimal path $\boldsymbol{q}^{\mathbf{*}}=q_{1}^{*},q_{2}^{*},...,q_{T}^{*}$ can be traced back as：

$\left\{ \begin{matrix} q_{t}^{*}=\underset{1\leq i\leq k}{\mathrm{argmax}} \delta_{t}(i) t=T \\ q_{t}^{*}=\psi_{t+1}(q_{t+1}^{*}) t=T-1, T-2,...,1 \end{matrix} \right.$	(10)

Finally, the transition pattern of IBSS is defined as the transition matrix of the hidden state time course, which refers to the likelihood of moving between each pair of state.

**IBSS-GAT Framework**

To adapt the individual-specific brain state space (IBSS) for downstream tasks, each participant's IBSS was treated as an individual graph (nodes: individual-specific brain states $\boldsymbol{\mu}=\left\{ \boldsymbol{\mu}_{\boldsymbol{1}},\boldsymbol{\mu}_{\boldsymbol{2}},\ldots,\boldsymbol{\mu}_{\boldsymbol{k}} \right\}\boldsymbol{,}\boldsymbol{\mu}_{\boldsymbol{i}}\in\mathbb{R}^{N}$; edges: transitions between states) and processed using a 2-layer (2 heads in the first, 1 in the second) Graph Attention Network (GAT) (4), which adaptively weighted and aggregated information from directly connected neighboring states within the graph. Specifically, given a shared weight matrix $\boldsymbol{W}\in\mathbb{R}^{F\times N}$, and a shared attentional mechanism $\boldsymbol{a}\in\mathbb{R}^{2F}$, the attention coefficient between connected states with transition from $s_{j}$ to $s_{i}$ is calculated as

$e_{ij}={\mathrm{LeakyReLU}\boldsymbol{(a}}^{\boldsymbol{T}}[\boldsymbol{W}\boldsymbol{\mu}_{\boldsymbol{i}}\parallel\boldsymbol{W}\boldsymbol{\mu}_{\boldsymbol{j}}])$(11)

For all neighboring states with direct transitions to $s_{i}$, we normalize the coefficients:

$\alpha_{ij}=\mathrm{softmax}_{j}\left( e_{ij} \right)=\frac{\exp\left( e_{ij} \right)}{\sum_{k\in\mathcal{N}_{i}} \exp\left( e_{ik} \right)}$	(12)

where $\mathcal{N}_{i}$ is the first-order neighborhood of state $s_{i}$ (including $s_{i}$). Thus, the aggregated features of $s_{i}$ are obtained as the linear combination of all neighbors weighted by the corresponding $\alpha_{ij}$:

$\boldsymbol{\mu}_{\boldsymbol{i}}^{\boldsymbol{'}}=\sum_{j\in\mathcal{N}_{i}} \alpha_{ij}\boldsymbol{W}\boldsymbol{\mu}_{\boldsymbol{j}}$	(13)

For the first layer with a multi-head attention mechanism, the aggregated features from each head are concatenated, yielding the following state embeddings:

$\boldsymbol{\mu}_{\boldsymbol{i}}^{\boldsymbol{'}}=\parallel_{c=1}^{C}\boldsymbol{\mu}_{\boldsymbol{i}}^{\boldsymbol{'(c)}}\boldsymbol{\in}\mathbb{R}^{CF}$	(14)

where $C$ is the number of attention heads. Finally, global mean pooling is applied to the output state embeddings of the final layer, averaging across all states to generate a fixed-dimensional, size-normalized global representation $\boldsymbol{\mu}^{\boldsymbol{out}}\boldsymbol{\in}\mathbb{R}^{CF}$ for each sample, which enables batch training and classification regardless of state number and identity.

**Model-derived cognitive signatures**

We used GNNExplainer (5) to compute the feature attributes underlying the class labels in the HCP-task cohort. This analysis yielded a measure of feature importance associated with the 8-cognitive-task-classification in each brain region for each sample (Figure S1). The derived feature attribution maps of each task were compared with that of rest using pairwise t-tests, and the spatial profile of brain regions with task-rest t-statistic difference (T value) >0 was mapped to canonical functional networks (17 instrinsic functional connectivity networks according to Yeo et al. (6) and 12 Cole-Anticevic networks (CAB-NP) defined by Ji (7), which complement each other, collectively covering all major networks implicated in cognitive processing) using the Network Correspondence Toolbox (NCT) (8) to identify subnetworks that discriminated between rest and each cognitive task (Figure S2). Specifically, for each functional network, spatial correspondence with the task-rest difference map was assessed as the Dice coefficient of network overlap, and spin tests with 1,000 permutations was used to quantify levels of significance (8).

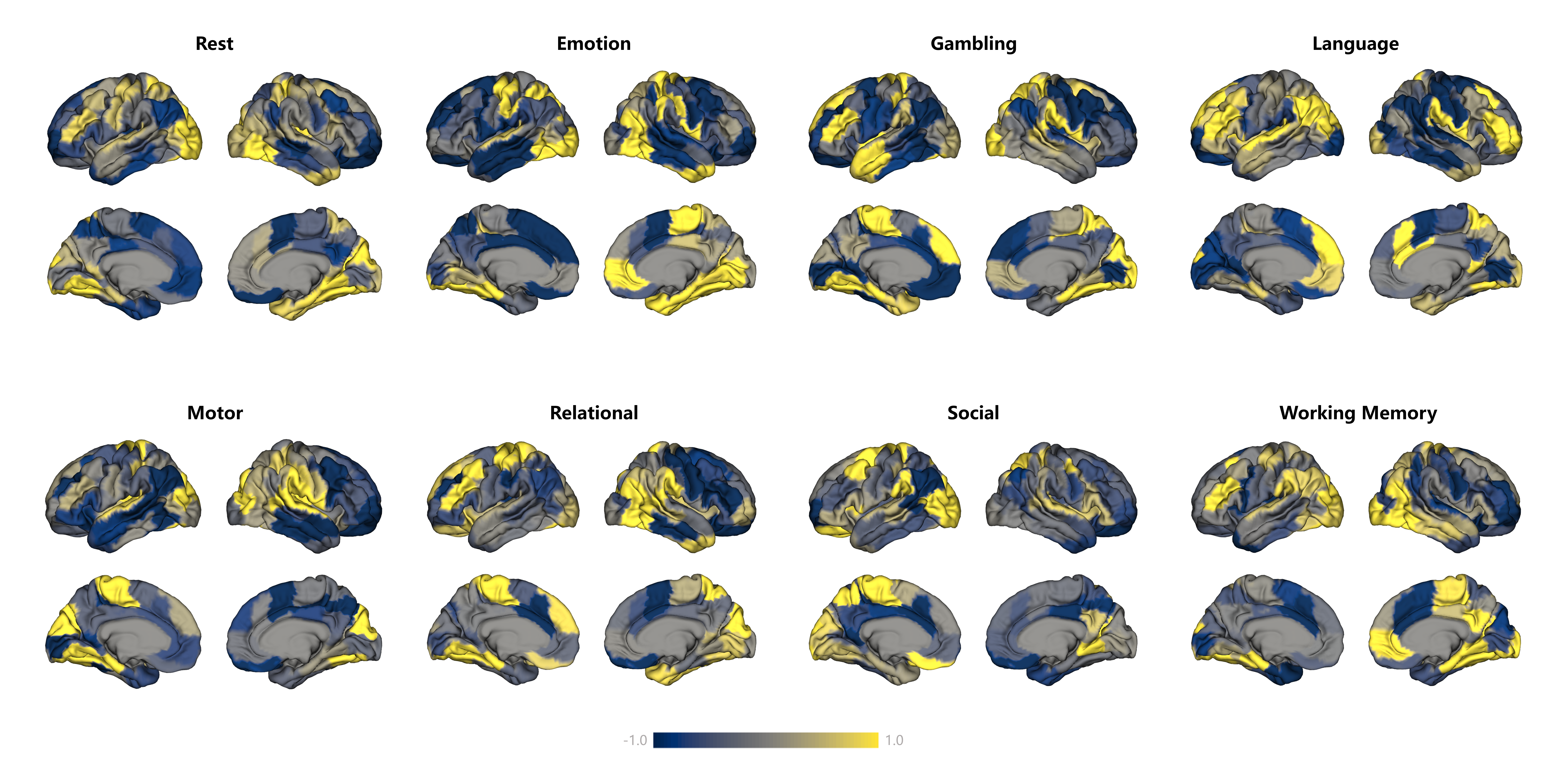

**Figure S1.** The feature attribution map for each task in cognitive decoding. The color bar indicates the average GNNExplainer-derived feature importance in each brain region.

In cognitive decoding, our model identified the dynamic properties associated with the SMN (Dice = 0.183, p_spin_ = 0.006), DAN (Dice = 0.174, p_spin_ = 0.010) and limbic system (Dice = 0.172, p_spin_ = 0.012) as key discriminative features of the emotion task. This corroborates prior evidence that emotional processing extends beyond isolated affective regions, requiring a coordinated network response where limbic-mediated salience detection is tightly coupled with top-down attentional appraisal and sensorimotor engagement (9-11).

With respect to the language task, our model highlighted the contribution of the FPCN (Dice = 0.312, p_spin_ = 0.002), CON (Dice = 0.280, p_spin_ = 0.001), and language areas (Dice = 0.134, p_spin_ = 0.048), which aligns with previous studies showing that language processing relies not only on perisylvian regions but also on domain-general control systems, where the FPCN supports flexible, context-dependent regulation of linguistic representations and the CON contributes to sustained task control and performance monitoring (12, 13).

Furthermore, our model delineated the DMN (Relational: Dice = 0.407, p_spin_ = 0.012; Social: Dice = 0.244, p_spin_ = 0.003; Working Memory: Dice = 0.384, p_spin_ = 0.012) and FPCN (Relational: Dice = 0.260, p_spin_ = 0.028; Social: Dice = 0.206, p_spin_ = 0.004; Working Memory: Dice = 0.310, p_spin_ = 0.008) as the common primary drivers distinguishing the relational, social, and working memory tasks. These tasks necessitate higher-order cognitive integration, particularly abstract reasoning, mental state inference, and the active manipulation of internal representations (14). The DMN is classically known to subserve internally directed processing, including self-referential thought and social cognition (15-17). In contrast, the FPCN orchestrates flexible cognitive control, dynamically adjudicating information flow between internally and externally directed systems (18, 19). Consequently, the observed DMN-FPCN coupling likely reflects a synergistic mechanism that integrates internally generated mental models with goal-directed executive control—an integration fundamental to complex cognition across these domains (20, 21).

Beyond the shared DMN–FPCN circuitry, our model delineated the salience/ventral attention network (SVAN-A: Dice = 0.142, p_spin_ = 0.037; SVAN-B: Dice = 0.147, p_spin_ = 0.045) and limbic system (Dice = 0.131, p_spin_ = 0.003) as distinct neural signatures of the social task. This unique network recruitment reflects the specialized neurobiological requirements of social cognition, which hinges on detecting salient social cues via the SVAN (22-24) and generating appropriate affective evaluations via the limbic system (25, 26). Consequently, unlike relational and working memory tasks, the social task necessitates a broader network topography that explicitly integrates emotion-laden, socially meaningful processing into higher-order cognitive frameworks (27).

Given that the core phenotypic features of ASD are deeply intertwined with performance deficits across these specific social and cognitive domains, it is highly plausible that ASD psychopathology is underpinned by aberrant dynamic interactions among these task-relevant networks. Most notably, the ubiquitous involvement of the FPCN—identified by our model as a key discriminative feature in four of the five tasks (excluding emotion)—suggests that its dysregulation may serve as a central neural locus for the broader executive and integrative dysfunctions characteristic of the disorder.

Besides, specific network features were also identified for the gambling and motor task: for gambling, IBSS-GAT highlighted the contribution of the salience / ventral attention network (SVAN-A, Dice = 0.181, p_spin_ = 0.006), which mediates salient stimuli detection and reward/punishment processing in addiction (28); while the somatomotor network (SM-B, Dice = 0.245, p_spin_ = 0.001) contributed the most to the identification of motor task. These results demonstrate that our model can faithfully capture biologically meaningful and cognitively aligned dynamic patterns.

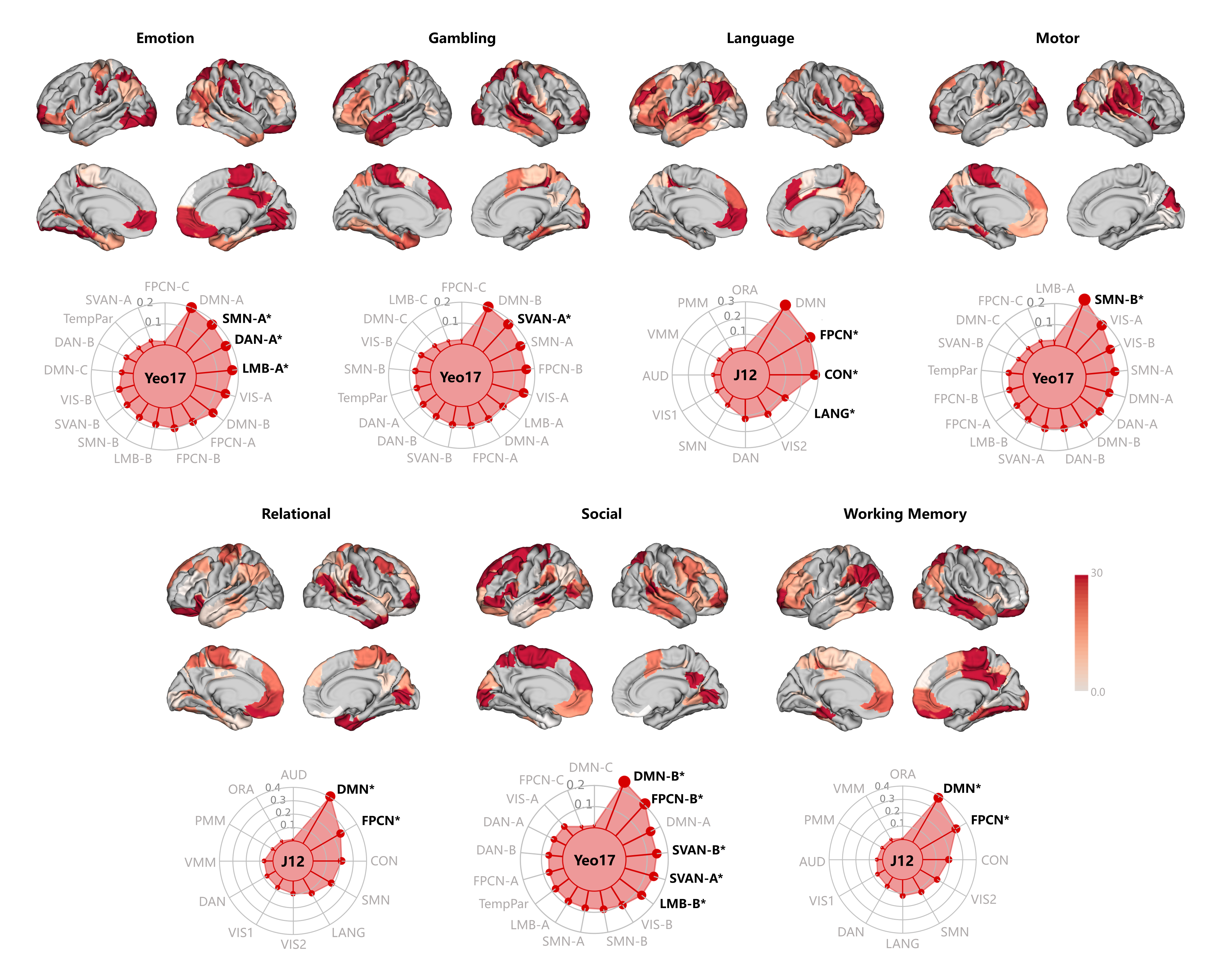

**Figure S2.** Model-derived cognitive signatures. The upper panels depict cortical surface projection of the feature attribution maps for 7 cognitive tasks. The color bar indicates the t-statistic difference (T value) of feature importance between the cognitive task and rest in each brain region. The lower panels show the spatial correspondence of the feature attribution maps with canonical functional networks. Each spot denotes the Dice coefficient of network overlap. *: Significant network overlap (p_spin_ < 0.05). SMN, somatomotor network; DAN, dorsal attention network; LMB, limbic system; SVAN, salience / ventral attention network; FPCN, frontoparietal control network; CON, cingulo-opercular network; LANG, language system; DMN, default mode network.

**Table S1. Parameters of task and rest scans in the HCP-task cohort**

| Task | Duration (s) | Response rate (s^-1^) | Block length (s) |
| --- | --- | --- | --- |
| Rest | 873 | 0 | -- |
| Emotion | 136 | 0.260 | 18 |
| Gambling | 192 | 0.172 | 28 |
| Language | 237 | 0.058 | 30 |
| Motor | 212 | 0.477 | 12 |
| Relational | 176 | 0.159 | 16 |
| Social | 207 | 0.025 | 23 |
| Working memory | 301 | 0.053 | 25 |

**Table S2. Demographic information for the ASD and TD groups in the ABIDE Cohorts**

|  | ABIDE-I | |  | ABIDE-II | |
| --- | --- | --- | --- | --- | --- |
|  | ASD (N = 436) | TD (N = 470) |  | ASD (N = 324) | TD (N = 369) |
| Age (years) | 17.32 ± 8.20 [7.00-64.00] | 17.10 ± 7.52 [6.47-56.20] |  | 14.28 ± 6.69 [5.22-54.00] | 14.16 ± 6.41 [5.89-37.00] |
| Sex (F/M) | 53/383 | 87/383 |  | 43/281 | 92/277 |
| FIQ | 106.08 ± 17.04 | 111.49 ± 12.67 |  | 107.00 ± 16.67 | 114.45 ± 12.54 |
| ADOS Social | 8.25 ± 2.78 | -- |  | -- | -- |
| ADOS Communication | 3.97 ± 1.53 | -- |  | -- | -- |
| ADOS Total | 12.22 ± 3.82 | -- |  | -- | -- |
| ADOS-2 Social Affect | -- | -- |  | 8.94 ± 3.76 | -- |
| ADOS-2 Total | -- | -- |  | 12.24 ± 4.41 | -- |

Note: All data are presented in mean ± standard deviation mode. In the ABIDE-I cohort, age, IQ and ADOS data were available for 906, 840, and 276 participants, respectively. In the ABIDE-II cohort, age, IQ and ADOS-2 data were available for 693, 614, and 143 participants, respectively. ASD, Autism Spectrum Disorder; TD, Typically Developing; FIQ, Full-scale Intelligence Quotient; ADOS, Autism Diagnostic Observation Schedule; ADOS-2, Autism Diagnostic Observation Schedule, Second Edition.

**Table S3. Demographic information for the MDD and HC groups in the DIRECT-II Cohort**

|  | DIRECT-II | |
| --- | --- | --- |
|  | MDD (N = 1283) | HC (N = 1118) |
| Age (years) | 33.55 ± 13.07 [11.00 - 77.00] | 33.17 ± 13.20 [13.00 – 67.00] |
| Sex (F/M) | 798 / 485 | 649 / 469 |
| Education | 12.04 ± 3.63 | 13.69 ± 3.55 |
| HAMD Total | 21.67 ± 7.24 | -- |
| HAMA Total | 18.37 ± 8.67 | -- |

Note: All data are presented in mean ± standard deviation mode. Age, Education and scale data (HAMD, HAMA) were available for 2401, 2401, and 726 participants, respectively. MDD, major depressive disorder; HC, healthy control; HAMD, Hamilton rating scale for depression; HAMA, Hamilton anxiety scale.

**Table S4. 10-fold cross-validation performance of IBSS-GAT**

| **Fold** | **HCP-task** | |  | **ABIDE-I** | | | |  | **DIRECT-II** | | | |
| --- | --- | --- | --- | --- | --- | --- | --- | --- | --- | --- | --- | --- |
|  | **ACC (%)** | **AUC (%)** |  | **ACC (%)** | **SEN (%)** | **SPE (%)** | **AUC (%)** |  | **ACC (%)** | **SEN (%)** | **SPE (%)** | **AUC (%)** |
| Fold1 | 99.58 | 99.97 |  | 81.32 | 75.00 | 87.23 | 79.45 |  | 70.12 | 75.97 | 63.39 | 68.64 |
| Fold2 | 98.73 | 99.96 |  | 80.22 | 77.27 | 82.98 | 86.17 |  | 72.92 | 66.41 | 80.36 | 74.61 |
| Fold3 | 99.58 | 99.99 |  | 76.92 | 77.27 | 76.60 | 79.69 |  | 71.67 | 71.09 | 72.32 | 72.66 |
| Fold4 | 98.94 | 99.91 |  | 68.13 | 54.55 | 80.85 | 71.08 |  | 68.33 | 71.88 | 64.29 | 73.28 |
| Fold5 | 99.36 | 99.94 |  | 81.32 | 79.55 | 82.98 | 82.21 |  | 71.25 | 72.66 | 69.64 | 75.11 |
| Fold6 | 99.36 | 99.98 |  | 78.02 | 81.82 | 74.47 | 74.76 |  | 73.33 | 80.47 | 65.18 | 77.78 |
| Fold7 | 98.52 | 99.85 |  | 78.89 | 81.40 | 76.60 | 81.59 |  | 71.25 | 78.12 | 63.39 | 75.45 |
| Fold8 | 99.58 | 99.99 |  | 76.67 | 62.79 | 89.36 | 78.97 |  | 70.42 | 78.91 | 60.71 | 72.80 |
| Fold9 | 100.0 | 100.0 |  | 76.67 | 67.44 | 85.11 | 71.20 |  | 68.75 | 66.67 | 71.17 | 70.67 |
| Fold10 | 99.36 | 99.99 |  | 74.44 | 62.79 | 85.11 | 80.75 |  | 68.75 | 70.54 | 66.67 | 71.47 |
| **Avg** | **99.30±0.42** | **99.96±0.05** |  | **77.26±3.70** | **71.99±8.95** | **82.13±4.68** | **78.59±4.60** |  | **70.68±1.64** | **73.27±4.69** | **67.71±5.50** | **73.25±2.50** |

**Table S5. Classification performance of IBSS-GAT in the ABIDE-I cohort using different atlases across a range of parcellation granularities**

| **Atlas** | **ACC (%)** | **SEN (%)** | **SPE (%)** | **F1 (%)** | **AUC (%)** |
| --- | --- | --- | --- | --- | --- |
| Schaefer100 | 77.26±3.70 | 71.99±8.95 | 82.13±4.68 | 78.98±2.83 | 78.59±4.60 |
| Schaefer200 | 76.78±3.04 | 77.67±5.90 | 75.96±9.80 | 76.22±2.06 | 76.92±4.08 |
| Schaefer400 | 76.52±1.77 | 76.37±6.37 | 76.64±6.53 | 75.48±2.10 | 80.75±2.73 |

Note: All data are presented in mean ± standard deviation mode.

**Table S6. Network overlaps of the ASD vs. TD t-statistic difference map of feature attribution in ABIDE-I**

| **Functional networks** | **Dice** | **p_spin_ value** |
| --- | --- | --- |
| Visual network | 0.159 | 0.810 |
| Somatomotor network | 0.247 | 0.206 |
| Dorsal attention network | 0.197 | 0.141 |
| Salience / ventral attention network | 0.100 | 0.974 |
| Limbic system | 0.072 | 0.921 |
| **Frontoparietal control network** | **0.266** | **0.006*** |
| **Default mode network** | **0.427** | **0.001*** |

Note: Significant network overlaps were bolded. *: p_spin_ < 0.05 with 1,000 permutations.

**Table S7. Network overlaps of the MDD vs. HC t-statistic difference map of feature attribution in DIRECT-II**

| **Functional networks** | **Dice** | **p_spin_ value** |
| --- | --- | --- |
| Visual network | 0.218 | 0.446 |
| Somatomotor network | 0.106 | 0.985 |
| **Dorsal attention network** | **0.244** | **0.002*** |
| Salience / ventral attention network | 0.127 | 0.926 |
| Limbic system | 0.043 | 0.982 |
| **Frontoparietal control network** | **0.309** | **0.001*** |
| **Default mode network** | **0.427** | **0.001*** |

Note: Significant network overlaps were bolded. *: p_spin_ < 0.05 with 1,000 permutations.

**Table S8. Network overlaps of the activation pattern of Metastate 1 in ABIDE-I**

| **Functional networks** | **Dice** | **p_spin_ value** |
| --- | --- | --- |
| Visual network | 0.011 | 0.990 |
| Somatomotor network | 0.188 | 0.593 |
| Dorsal attention network | 0.201 | 0.198 |
| **Salience / ventral attention network** | **0.302** | **0.001*** |
| Limbic system | 0.116 | 0.634 |
| **Frontoparietal control network** | **0.390** | **0.001*** |
| Default mode network | 0.350 | 0.301 |

Note: Significant network overlaps were bolded. *: p_spin_ < 0.05 with 1,000 permutations.

**Table S9. Network overlaps of the activation pattern of Metastate 3 in DIRECT-II**

| **Functional networks** | **Dice** | **p_spin_ value** |
| --- | --- | --- |
| Visual network | 0 | 0.986 |
| Somatomotor network | 0.334 | 0.052 |
| Dorsal attention network | 0.026 | 0.995 |
| **Salience / ventral attention network** | **0.258** | **0.020*** |
| Limbic system | 0.247 | 0.152 |
| Frontoparietal control network | 0.278 | 0.222 |
| **Default mode network** | **0.421** | **0.038*** |

Note: Significant network overlaps were bolded. *: p_spin_ < 0.05 with 1,000 permutations.

**Table S10. Network overlaps of the activation pattern of Metastate 4 in DIRECT-II**

| **Functional networks** | **Dice** | **p_spin_ value** |
| --- | --- | --- |
| Visual network | 0.048 | 0.911 |
| Somatomotor network | 0.024 | 0.999 |
| Dorsal attention network | 0.018 | 0.998 |
| Salience / ventral attention network | 0.017 | 1 |
| **Limbic system** | **0.361** | **0.001*** |
| Frontoparietal control network | 0.246 | 0.097 |
| **Default mode network** | **0.650** | **0.001*** |

Note: Significant network overlaps were bolded. *: p_spin_ < 0.05 with 1,000 permutations.

**Table S11. Network overlaps of the ASD vs. TD t-statistic difference map of feature attribution in ABIDE-II**

| **Functional networks** | **Dice** | **p_spin_ value** |
| --- | --- | --- |
| Visual network | 0.244 | 0.284 |
| Somatomotor network | 0.196 | 0.615 |
| Dorsal attention network | 0.160 | 0.452 |
| Salience / ventral attention network | 0.149 | 0.651 |
| Limbic system | 0.144 | 0.419 |
| **Frontoparietal control network** | **0.262** | **0.022*** |
| **Default mode network** | **0.345** | **0.045*** |

Note: Significant network overlaps were bolded. *: p_spin_ < 0.05 with 1,000 permutations.

**Table S12. Network overlaps of the FPCN-dominated metastate underlying IBSS-derived neural representations in ABIDE-II**

| **Functional networks** | **Dice** | **p_spin_ value** |
| --- | --- | --- |
| Visual network | 0.314 | 0.193 |
| Somatomotor network | 0.033 | 0.984 |
| Dorsal attention network | 0.077 | 0.894 |
| Salience / ventral attention network | 0.213 | 0.078 |
| Limbic system | 0.145 | 0.286 |
| **Frontoparietal control network** | **0.349** | **0.005*** |
| Default mode network | 0.307 | 0.240 |

Note: Significant network overlaps were bolded. *: p_spin_ < 0.05 with 1,000 permutations.

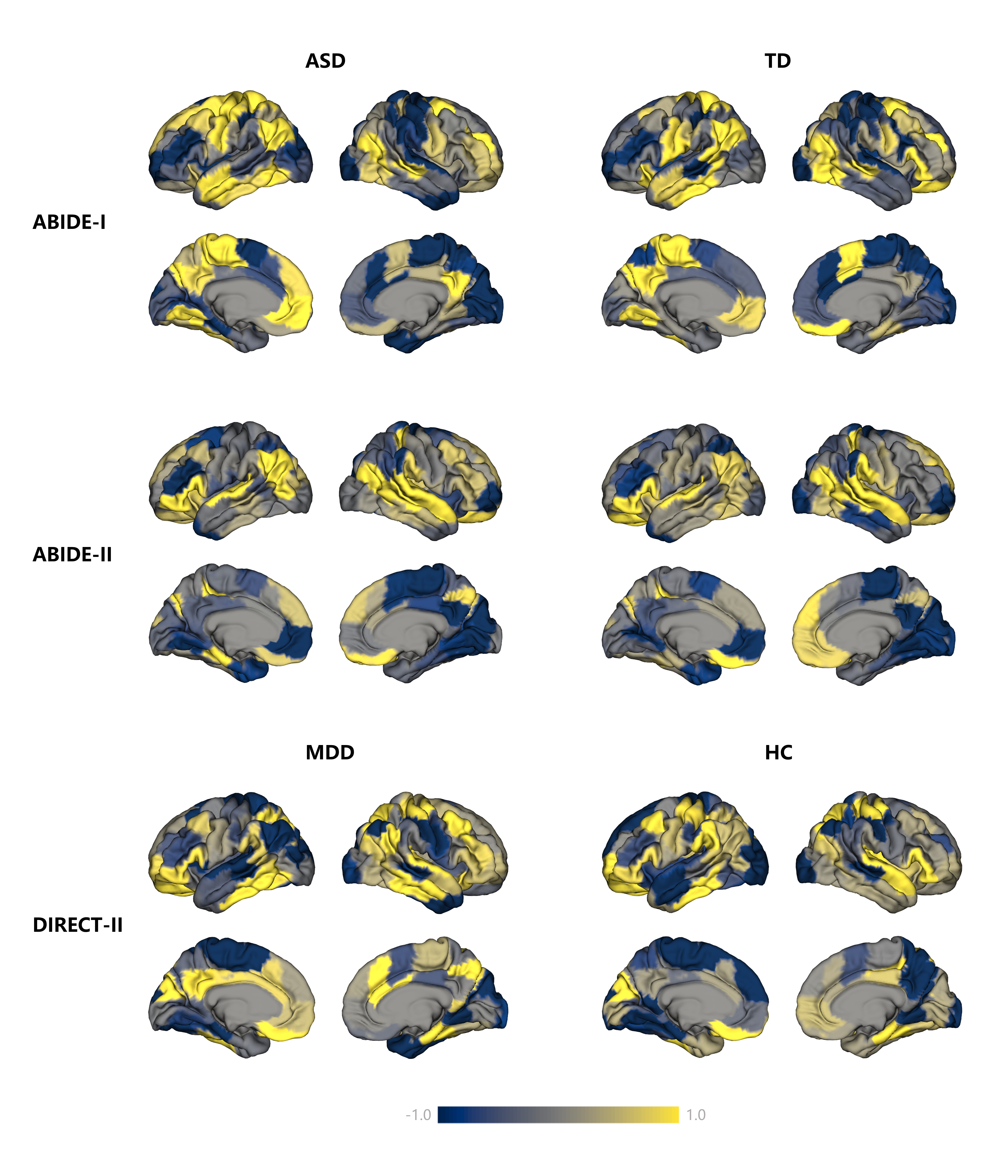

**Figure S3.** Zero-centered feature attribution maps for the ASD and TD groups in the ABIDE-I and ABIDE-II cohorts, and for the MDD and HC groups in the DIRECT-II cohort. The color bar indicates the group-average GNNExplainer-derived feature importance in each brain region.

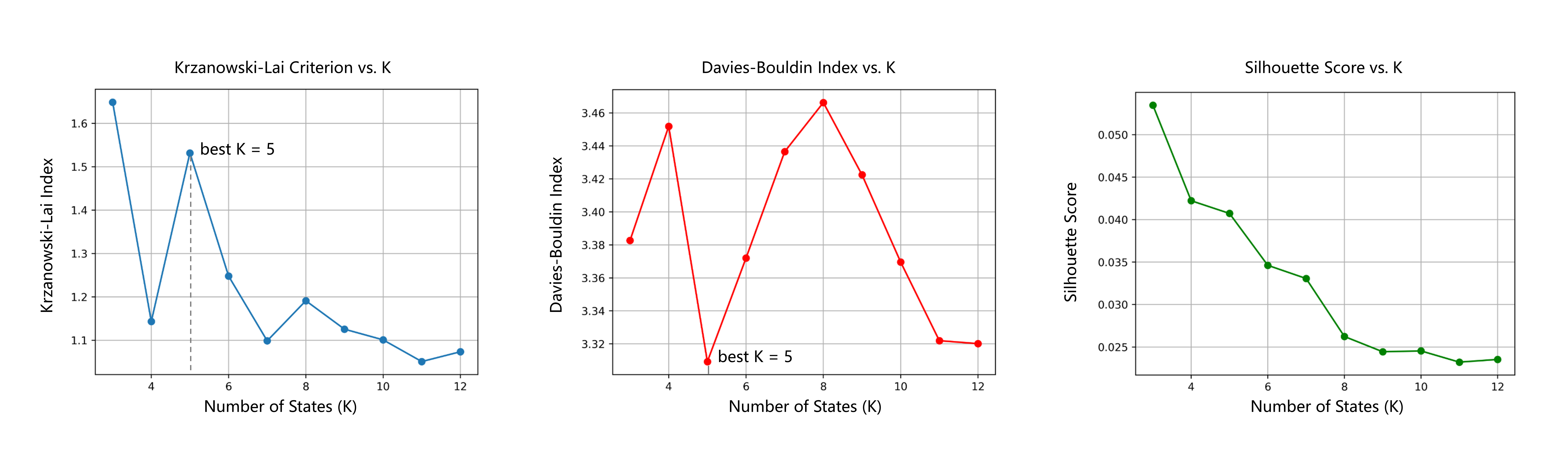

**Figure S4.** Determination of the optimal number of metastates. The line plots illustrate how three clustering metrics (Krzanowski-Lai criterion, Davies-Bouldin index, and Silhouette score) vary with the number of metastates.

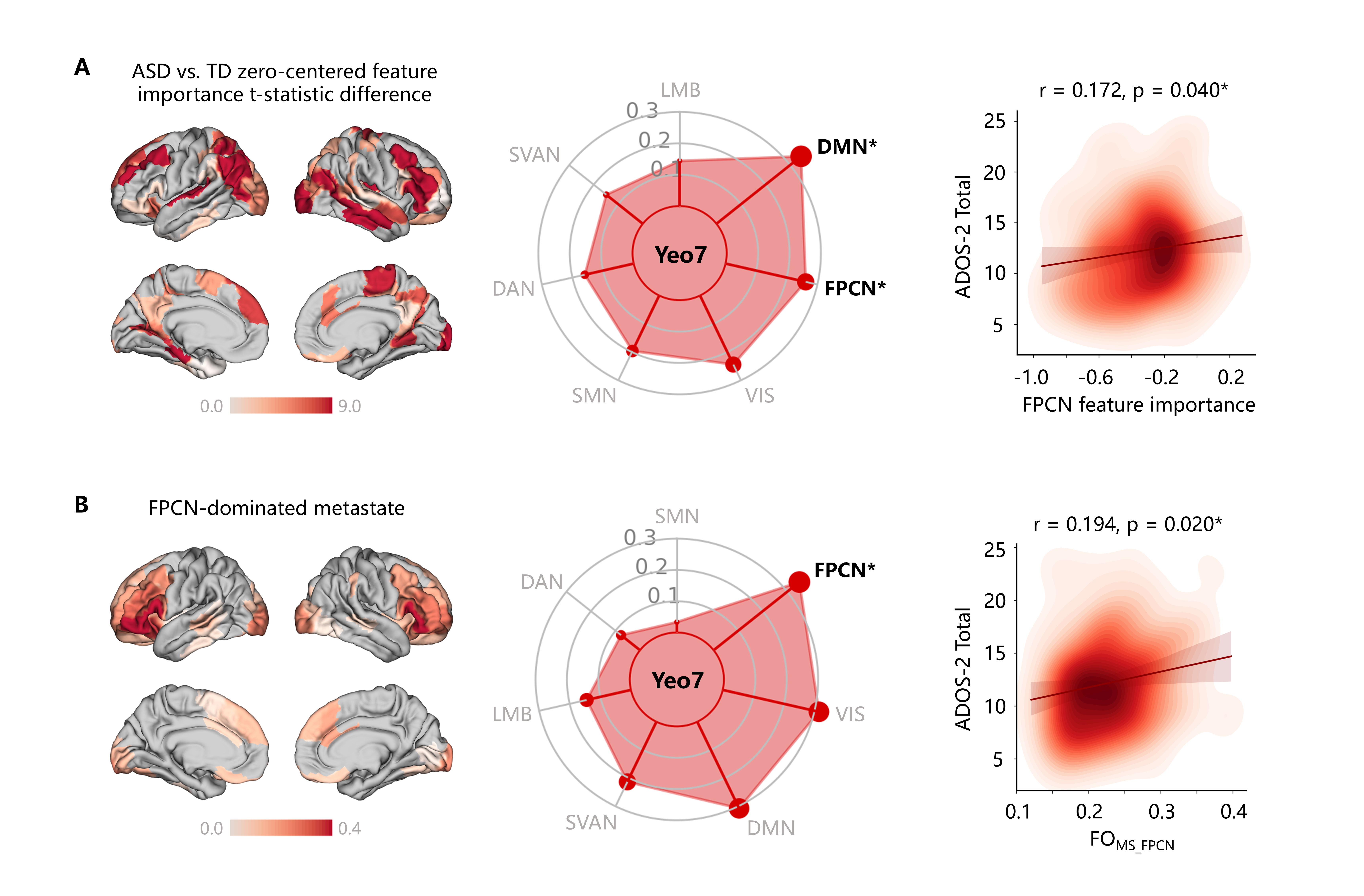

**Figure S5.** Independent replication of predictive and metastate-based interpretations in ABIDE-II. **(A)** Cortical maps show regions with greater GNNExplainer-derived, zero-centered feature importance in ASD than in TD, as indicated by positive t statistics. The radar plot summarizes their overlap with the Yeo seven-network parcellation. FPCN feature importance was positively associated with ADOS-2 total scores in individuals with ASD. **(B)** Activation pattern and network overlap of the independently reconstructed FPCN-dominated metastate. Its fractional occupancy was positively associated with ADOS-2 total scores. Density contours illustrate the joint distributions of neural measures and clinical scores; lines and shaded bands indicate fitted trends and 95% confidence intervals. Correlations were assessed using Spearman’s rank correlation. Asterisks indicate significant network overlap or clinical associations. ASD, autism spectrum disorder; TD, typically developing; FPCN, frontoparietal control network; FO, fractional occupancy; ADOS-2, Autism Diagnostic Observation Schedule, Second Edition.
